# Quality Appraisal of Screening Guidelines for Cardiovascular disease Prevention: *a scoping review*

**DOI:** 10.1101/2023.08.05.23293686

**Authors:** Mohammed Abd ElFattah Mohammed Darwesh Badawy, Lin Naing, Iman Ahmad Saeed, Nik Ani Afiqah Tuah

## Abstract

**Objective:** Each nation develops a guideline for managing and preventing cardiovascular disease (CVD) based on the available literature and the evidence-based consensus in their guideline’s recommendations. The goal of this review is to appraise the quality of many guidelines recommended by several international organizations for the early detection and prevention of CVD.

**Methods:** We conducted a scoping review using a few guideline-specific databases using Preferred Reporting Items for Systematic Reviews and Meta-Analyses extension for Scoping Reviews (PRISMA-ScR) as the reporting guideline for this review. We used the Appraisal of Guidelines for Research and Evaluation II (AGREE II) instrument to rate the methodological quality and rigorousness of the included guidelines for CVD prevention and early detection.

**Results:** Out of the 2,466 guidelines identified by our search, based on our eligibility criteria, we included a total of 20 distinct guidelines globally focused on screening for CVD prevention and early detection. (**70%**) of these guidelines were classified as "strongly recommended" while only (**90%**) were classified as just "recommended". The AGREE II appraisal domains with the highest median scores (percentile 25^th^, 75^th^) were “Clarity of Presentation” **92**% (81, 95.5), followed by Applicability **88**% (73, 93). The Rigour of Development and Editorial Independence were the lowest scores with a median of **78**% (66.5, 84) and **75**% (60.5, 92).

**Conclusions:** Using AGREE II quality appraisal tool, we provided a critical quality appraisal of several international CVD prevention guidelines and reveal opportunities to improve the CVD prevention guidelines’ quality.

**What is already known in this review?:**

- Each community develops a guideline for screening and managing CVD prevention based on the current evidence and evidence-based consensus.
- AGREE II tool is an internationally standardized instrument to report the rigorousness of development, transparency, and methodological quality of the clinical practice guidelines.

**What this review adds:**

- This review provided a critical quality appraisal of several international CVD prevention guidelines.
- The highest AGREE II domain median score was “Clarity of Presentation” while the least domains were “Editorial Independence” and “Rigour of Development”.
- **70%** of these guidelines were classified as "strongly recommended".

**How this review might affect research, practice, or policy:**

- This review provided a critical quality appraisal of many CVD prevention guidelines and revealed opportunities to improve CVD prevention guidelines’ quality.

## INTRODUCTION

Cardiovascular disease (CVD) is the leading cause of disability and premature death around the world, and contributes substantially to the escalating health care costs as the prevalence of CVD is estimated to reach 130 million patients with related mortality of about 24 million deaths by 2035 that could incur health care sector globally costs more than trillion dollars [1,2]. CVD is defined as any illness that affects the heart or the blood vessels, including, for example, coronary artery disease (CAD), cerebrovascular disease, peripheral arterial disease, other heart diseases and deep vein thrombosis [3]. The main pathology in CVD is atherosclerosis, which develops and advances with age and usually, the symptoms occur late in middle age commonly in the form of acute coronary and cerebrovascular events which happen suddenly and are often fatal before seeking the appropriate medical support [4].

Cardiovascular disease (CVD) prevention refers to a systematic plan of actions aiming at reducing and eliminating the disabilities related to CVDs and these actions could be targeted at the population level or an individual level [5]. The ultimate goal of CVD prevention is the reduction of CVD prevalence and subsequently CVD-related deaths worldwide so the proper and thorough implementation of the recommended guideline for CVD preventive measures is of ultimate concern for all national organizations and joint societies of the healthcare sector [6,7].

Population-targeted prevention strategies emphasize decreasing the overall exposure of the population to CVD risk factors by modifying the lifestyle regardless of the individual CVD risk. However, Individual-based prevention strategies are targeted at high-risk groups to prevent the onset of CVD through individual risk factors’ reduction [8]. CVD risk means the risk of suffering fatal or nonfatal CVD events, for instance, the risk of myocardial infarction or stroke in the upcoming years [4]. Individual CVD risk prevention includes the ‘vertical’ approach which aims at managing a single risk according to predefined cut-offs regardless of the presence of concomitant risk factors or the ’total’ cardiovascular risk approach in preventing CVD which depends on the individual’s odds of having fatal or nonfatal CVD events in a predetermined period with regard of the presence of several related risk factors rather than single risk factor [9].

Each country or joint society develops a guideline for screening and managing CVD prevention based on the current evidence and identifying remaining knowledge gaps by appraising their evidence-based consensus in their guidelines [5]. Several approaches to CVD risk management can prevent fatal and non-fatal CVD events and decisions about recommending a specific approach for the prevention of CVD should be guided by an estimation of the risk of any such CVD event [4].

There are several risk-prediction charts and online risk calculators that accompany these guidelines allowing management to be targeted according to simple risk calculations of the predicted CVD event. Several risk calculators are developed to estimate the individuals’ total CVD events risk or to directly assess one of the major CVD event risks, such as Stroke Riskometer™, a unique tool for assessing the specific risk of Stroke and endorsed by the World Stroke Organization [10]. Every national healthcare system should use a CVD risk calculator that is more feasible, precise, and user-friendly and is tailored to the population based on several important features, including variables, predictive accuracy, discrimination index, applicability, understandability, and cost-effectiveness [11].

Recommendations are formulated in these guidelines by different health organizations and societies for the management of major CVD risk factors and preventing CVD events risk through a set of lifestyle modification advice, a protocol for specific screening tests and many prophylactic drug therapies targeted to each CVD risk category. These guidelines provide an endorsed framework for the development of national guidance on the prevention of CVD risk in their population taking into account the particular political, economic, social and medical circumstances [4]. However, many of them are not even supported by solid scientific evidence. Additionally, not all of them harness techniques for evaluating the quality of a body of evidence and the strength of recommendations formulating the guideline framework [12].

The goal of this scoping review is to provide an overview and evaluation of the methodological rigour and quality of the many CVD screening guidelines that have been published internationally and advocated for CVD prevention and early detection by independent organizations, societies, and health authorities.

## METHODS OF SCOPING REVIEW

### Study and search strategy

We conducted a scoping review searching for cardiovascular disease screening guidelines using guideline-specific databases such as Guidelines International Network International Guideline Library, National Guideline Clearinghouse (United States), National Library for Health Guidelines Finder (United Kingdom) and Canadian Medical Association Clinical Practice Guidelines InfoBase. The duration of the search was from 2000 to 2022 and the guidelines written only in the English language were included in the results. The main keywords used include “cardiovascular disease”, “CVD prevention”, “cardiovascular disease prevention”, “cardiovascular disease screening”, “CVD screening”, “CVD screening guideline”, “cardiovascular risk screening”, “screening guideline” and “guideline”.

### Study selection

We used Preferred Reporting Items for Systematic reviews and Meta-Analyses extension for Scoping Reviews (PRISMA-ScR) in this review (**Figure 1**) [13], and filled the specific checklist for reporting scoping reviews (**Appendix 1**. PRISMA-ScR checklist). The initial literature search of this review yielded about 2,466 guidelines. We excluded all duplicates, parts of a guideline, guidelines written by an unknown organization, commentaries, guidelines not focused on CVD and other irrelevant guidelines. We reviewed 40 full-text guidelines and excluded the guidelines that are limited to a single CVD condition or not focused on screening pathways and only included 20 guidelines that met the eligibility criteria of this review.

**Figure 1.**
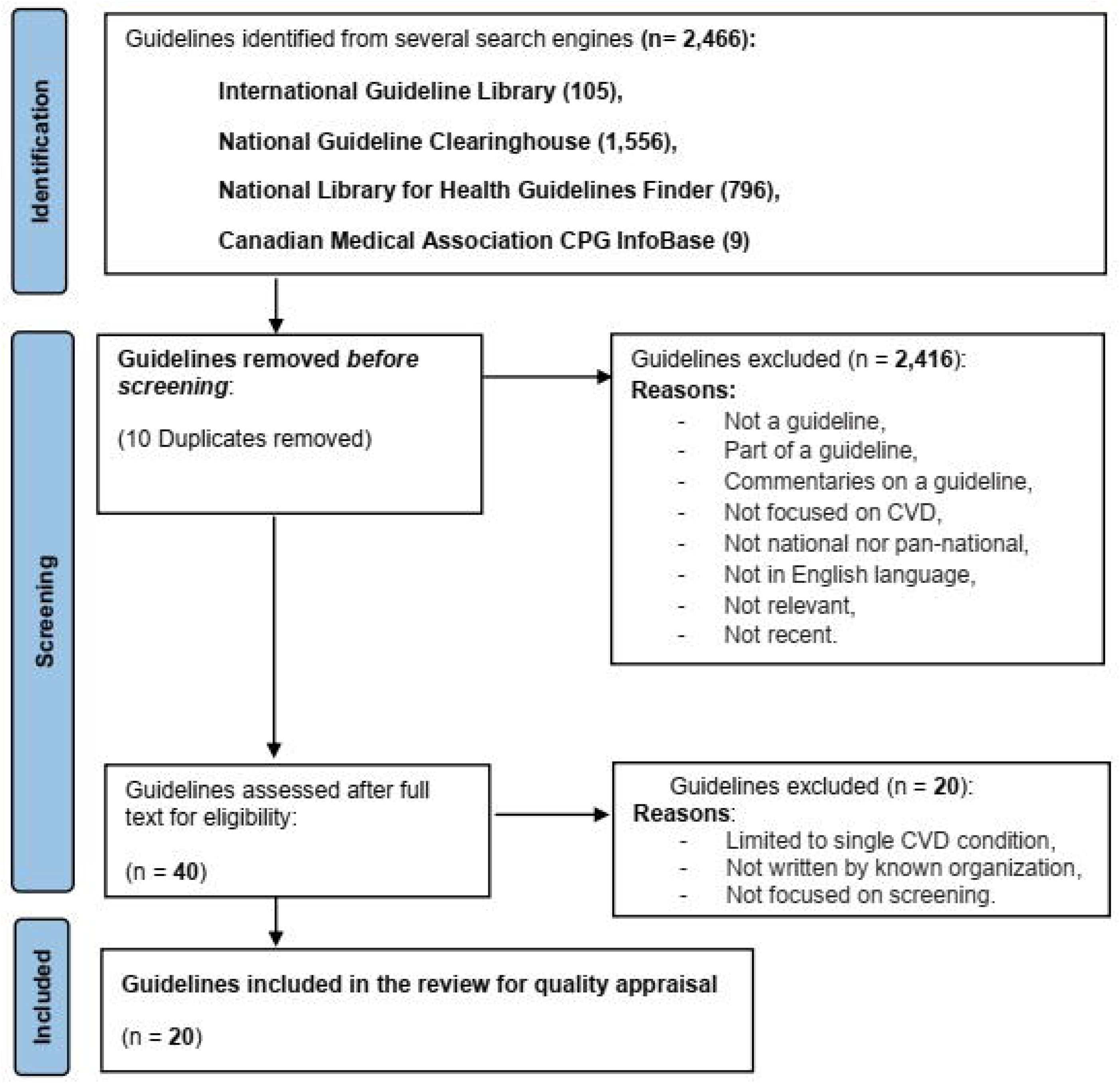
Identification of guidelines via databases.

### Guidelines’ quality appraisal

In this review, we used The ***A****ppraisal of **G**uidelines for **Re**search & **E**valuation (AGREE II)* instrument as an assessment tool for quality appraisal of the included screening guidelines for CVD prevention and early detection. The 23-item AGREE II tool is an internationally standardized instrument to report the rigorousness of development, transparency and methodological quality of the clinical practice guidelines [14,15]. A Pair of reviewers independently fulfil AGREE Reporting Checklist [16] (**Appendix 2**) with comments (**Appendix 3**) for every CVD screening guideline which is a checklist based on the AGREE II instrument via the online platform MYAGREE PLUS to ensure the comprehensive and independent appraisal of the 23 items organized into the six domains of AGREE II tool (Scope and Purpose, Stakeholder Involvement, Rigour of Development, Applicability, Clarity of Presentation and Editorial Independence).

A scale of 1 to 7 points, with 1 denoting "Strongly disagree," and 7 denoting "Strongly agree," was used to score each item. The sum was displayed as a percentage of the domain’s highest quality score (from 0 to 100 %). Upon completing that, the overall assessment is performed to make a judgment about the quality of each CVD screening guideline, rating the guideline quality from 1 (lowest quality) to 7 (highest quality) considering the appraisal items considered in the assessment process. Discrepancies were resolved by a consensus between two raters. Finally, the guideline was defined as “strongly recommended” if most domains (>3 domains) scored >60%; the guideline was “recommended,” if most domains scored between 30% and 60%, and if most domain scores were <30%, the guideline was “not recommended” [17].

## RESULTS

**Table 1** provides a detailed description of the characteristics of included 20 screening guidelines for CVD prevention and early detection entailing the guideline title, the year of release, the organizations incorporate in the development and appraisal of such guidelines, the geographic boundaries best to apply this clinical practice guideline and also key features that characterize each of these guidelines. Five of them were developed in North America (four in the United States of America and one in Canada), eight in Europe (two continental, two in Scotland, and four in the UK), Three in Asia (one each in Singapore, Malaysia, and Brunei Darussalam), two in Australia, one in New Zealand, and one were global collaborations.

**Table 1.** Characteristics of included screening guidelines for CVD prevention.

| Acronym /year | Guideline name | The organization developed the guideline | Geographic boundaries applied | Key features |
| --- | --- | --- | --- | --- |
| **JBS 2 [18] / 2005** | JBS 2: Joint British Societies' Guidelines On Prevention of Cardiovascular Disease in Clinical Practice | British Cardiac Society<br>British Hypertension Society<br>Diabetes UK<br>HEART UK<br>Primary Care Cardiovascular Society<br>The Stroke Association | United Kingdom | Targeted population with high total CVD risk.<br>The outcome risk is to develop a CVD event over 10 years.<br>Assess risk by **JBS CVD risk prediction chart**.<br>No evidence grading system was adopted. |
| **SHAPE [19] / 2006** | SHAPE (Screening for Heart Attack Prevention and Education) Guideline | Association for Eradication of Heart Attack | United States | Noninvasive screening of all asymptomatic population for risk of subclinical atherosclerosis using **Atherosclerosis test**.<br>No evidence grading system was adopted. |
| **WHO [4] / 2007** | Guidelines for assessment and management of cardiovascular risk | World Health Organization (WHO) | Worldwide | Targeted asymptomatic population with high total CVD and patients with symptoms of atherosclerosis.<br>The outcome risk is fatal and non-fatal CVD events over 10 years.<br>Assess risk by **WHO/ISH CVD risk charts**.<br>SIGN and GRADE evidence grading system adopted. |
| **SIGN [20] / 2007** | Risk estimation and the prevention of cardiovascular disease | Scottish Intercollegiate Guidelines Network (SIGN) | Scotland | Targeted asymptomatic population above 40 years.<br>The outcome risk is the **ASSIGN score**.<br>Assess risk by **The ASSIGN score online calculator**.<br>**SIGN** evidence grading system adopted. |
| **ACCF/AHA [21] / 2010** | Guideline for Assessment of Cardiovascular Risk in Asymptomatic Adults (ACCF/AHA Guideline) | The American College of Cardiology Foundation and the American Heart Association | United States | Targeted asymptomatic population above 20 years excluding those with known CVD disease.<br>The outcome risk is the 10-year risk of CHD.<br>Assess risk by Framingham Risk Score (**FRS**).<br>**AHA/ACC** evidence grading system adopted. |
| **NHS / 2011** | Primary Prevention of Cardiovascular Disease | National Health Services (NHS) | United Kingdom | Targeted population with high total CVD risk.<br>The outcome risk is the **ASSIGN score**.<br>Assess risk by **The ASSIGN score online calculator**.<br>**SIGN** evidence grading system adopted. |
| **MOH Singapore [22] / 2011** | Screening for cardiovascular disease and risk factors – MOH clinical practice guidelines | Ministry of Health – Singapore<br>Singapore Heart Foundation | Singapore | Targeted all Singaporean asymptomatic population above 20 years.<br>The outcome risk is the 10-year risk of CHD.<br>Assess risk by **FRS-based NCEP ATP III**.<br>**SIGN** evidence grading system adopted. |
| **NVDPA [23] / 2012** | Guidelines for the management of Absolute cardiovascular disease risk | **The National Vascular Disease Prevention Alliance (NVDPA):**<br>Diabetes Australia, the National Heart Foundation of Australia, Kidney Health Australia, the National Stroke Foundation | Australia | Targeted all Australian population aged 45–74 years.<br>The outcome risk is to develop a CVD event **over 5 years**.<br>Assess risk with the **Australian Absolute CVD Risk Calculator**.<br>**NHMRC** evidence grading system adopted. |
| **NICE [24] / 2014** | Cardiovascular disease: risk assessment and reduction, including lipid modification | National Institute for Health and Care Excellence (NICE) | United Kingdom | Targeted adults who are at risk or have CVD.<br>The outcome risk is the 10-year **QRISK<sup>®2</sup>** score.<br>& **QRISK<sup>®</sup> Heart Age**.<br>Assess risk by **QRISK2** risk assessment tool.<br>**NICE** evidence grading system adopted. |
| **JBS 3 [25] / 2014** | JBS 3: Joint British Societies' Guidelines On Prevention of Cardiovascular Disease in Clinical Practice | British Cardiac Society<br>British Hypertension Society<br>Diabetes UK<br>HEART UK<br>Primary Care Cardiovascular Society<br>The Stroke Association | United Kingdom | Targeted population with high total CVD risk.<br>Outcome risk is CVD event over 10 years and **the lifetime (Heart Age)**<br>Assess risk by **JBS3 risk online calculator**.<br>No evidence grading system was adopted. |
| **ADA [26] / 2015** | Cardiovascular Disease and Risk Management | American Diabetes Association | United States | Targeted 10-year CHD risk among U.S. adults with diabetes. |
| **ESC [27] / 2016** | European Guidelines on cardiovascular disease prevention in clinical practice | The Sixth Joint Task Force of the European Society of Cardiology | Europe | Targeted asymptomatic population > 40 years.<br>The outcome risk is only fatal CVD events over 10 years.<br>Assess risk by **SCORE risk charts**.<br>**ESC** evidence grading system adopted. |
| **SIGN [28] / 2017** | Risk estimation and the prevention of cardiovascular disease | Scottish Intercollegiate Guidelines Network (SIGN) | Scotland | Targeted population with a high risk of CVD events.<br>The outcome risk is the **ASSIGN score**. |
|  |  |  |  | Assess risk by **The ASSIGN score online calculator**.<br>**SIGN** evidence grading system adopted. |
| **MOH Malaysia**<br>[29]<br>/ 2017 | Primary & Secondary Prevention of Cardiovascular Disease | Ministry of Health Malaysia<br>Academy of Medicine Malaysia<br>National Heart Association of Malaysia | Malaysia | Targeted all individuals in Malaysia.<br>The outcome risk is the 10-year risk of CHD.<br>Assess risk by Framingham Risk Score (**FRS**).<br>**ESC** evidence grading system adopted. |
| **C-CHANGE**<br>[30]<br>/ 2018 | Canadian Cardiovascular Harmonized National Guidelines Endeavour (C-CHANGE) guideline for the prevention and management of cardiovascular disease in primary care | Canadian Cardiovascular Society<br>Canadian Association of Cardiovascular Prevention and Rehabilitation<br>Diabetes Canada<br>Hypertension Canada<br>Heart and Stroke Foundation | Canada | Targeted Canadian population with a high risk of CVD events.<br>The outcome risk is the 10-year risk of CHD.<br>Assess risk by modified **FRS**.<br>**GRADE** evidence grading system adopted. |
| **MOH New Zealand**<br>[31]<br>/ 2018 | Cardiovascular Disease Risk Assessment and Management for Primary Care | Ministry of Health New Zealand | New Zealand | The targeted population of New Zealand, all men aged 45–74 years, and all women aged 55–74 years.<br>The outcome risk is to develop a CVD event **over 5 years**.<br>Assess risk by **PREDICT** CVDRA equations.<br>No evidence grading system was adopted. |
| **AHA/ACC**<br>[32]<br>/ 2019 | AHA/ACC Guideline on the Primary Prevention of Cardiovascular Disease | The American Heart Association and The American College of Cardiology | United States | Targeted asymptomatic population aged 40 to 75 years.<br>Outcome risk is the 10-year risk for patients aged 40-79 or Lifetime risk for patients aged 20-59.<br>Assess risk by **ASCVD Risk Estimator**.<br>**AHA/ACC** evidence grading system adopted. |
| **NCDs BN**<br>[33]<br>/ 2019 | National Health Screening Guideline on Noncommunicable Diseases (NCDs) | Noncommunicable Diseases (NCDs) Prevention Unit, Ministry of Health, Brunei Darussalam | Brunei Darussalam | Targeted asymptomatic adult population 40 years and above.<br>The outcome risk is fatal and non-fatal CVD events over 10 years.<br>Assess risk by **WHO/ISH CVD risk prediction chart for WPR-A**<br>**NICE** evidence grading system adopted. |
| **NVDPA**<br>/ 2020 | Guidelines for the management of Absolute cardiovascular disease risk (Update) | **The National Vascular Disease Prevention Alliance (NVDPA):**<br>Diabetes Australia, the National Heart Foundation of Australia, Kidney Health Australia, the National Stroke Foundation | Australia | Targeted all Australian population aged 45–74 years.<br>The outcome risk is a CVD event **over 5 years**.<br>Assess risk with the **Australian Absolute CVD Risk Calculator**.<br>**NHMRC** evidence grading system adopted. |
| **ESC** [34]<br>/ 2021 | ESC Guidelines on cardiovascular disease prevention in clinical | Task Force for cardiovascular disease prevention in clinical practice | Europe | Targeted asymptomatic population aged 40-69 years.<br>The outcome risk is only fatal and |
|  | practice | European Society of Cardiology<br>European Association of Preventive Cardiology (EAPC) |  | CVD events over 10 years.<br>Assess risk by **SCORE2** & **SCORE2-OP**<br>**ESC** evidence grading system adopted. |

According to **Table 2**, the AGREE II appraisal domains with the highest median scores (percentile 25^th^, 75^th^) were Clarity of Presentation **92**% (81, 95.5), followed by Applicability **88**% (73, 93). While “Scope and Purpose”, and “Stakeholder Involvement”, were of intermediate rates and scored **83**% (78, 93) and **81**% (73.5, 90.5) respectively. The Rigour of Development and Editorial Independence were the lowest scores with a median of **78**% (66.5, 84) and **75**% (60.5, 92) respectively.

**Table 2.**
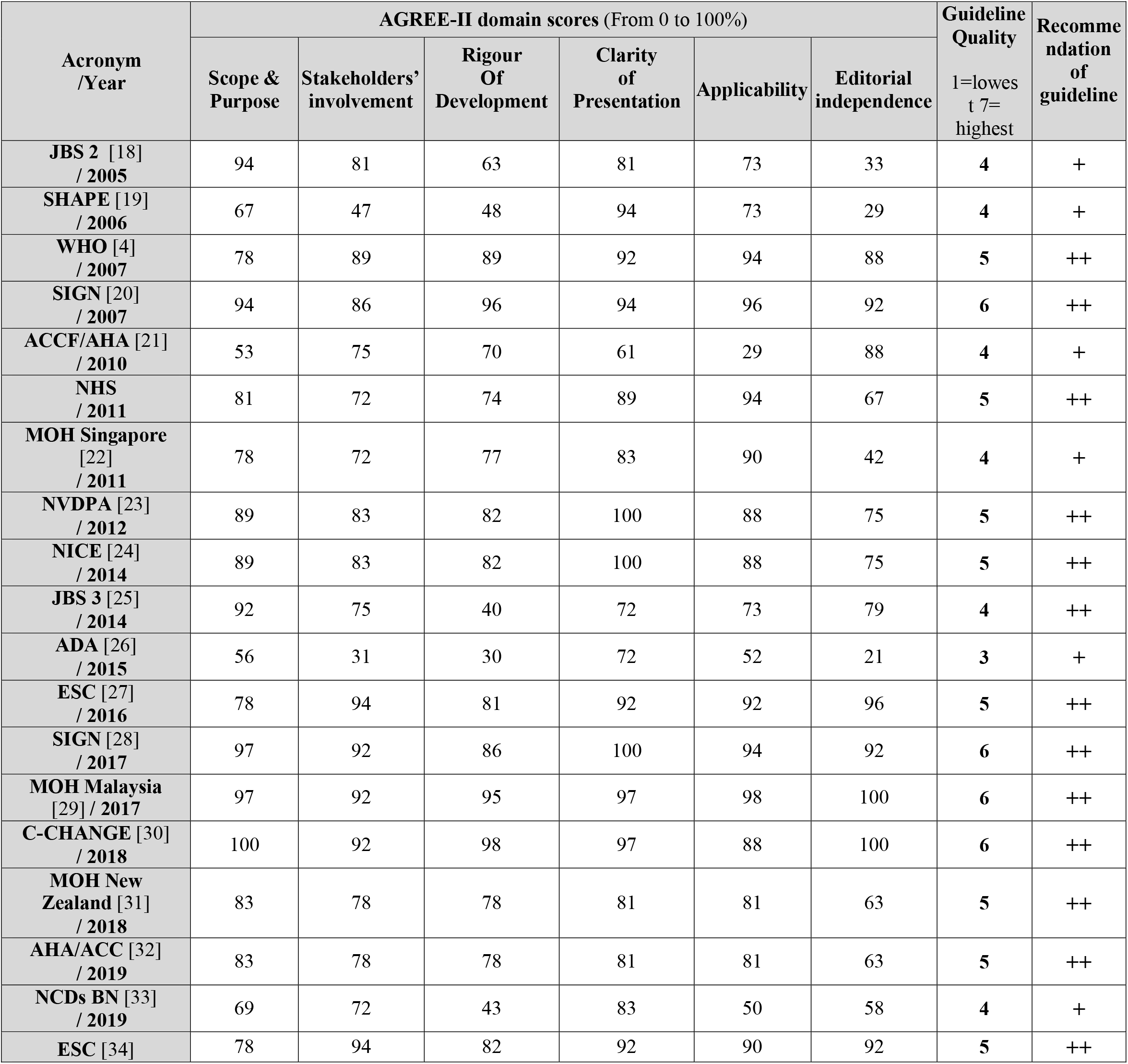
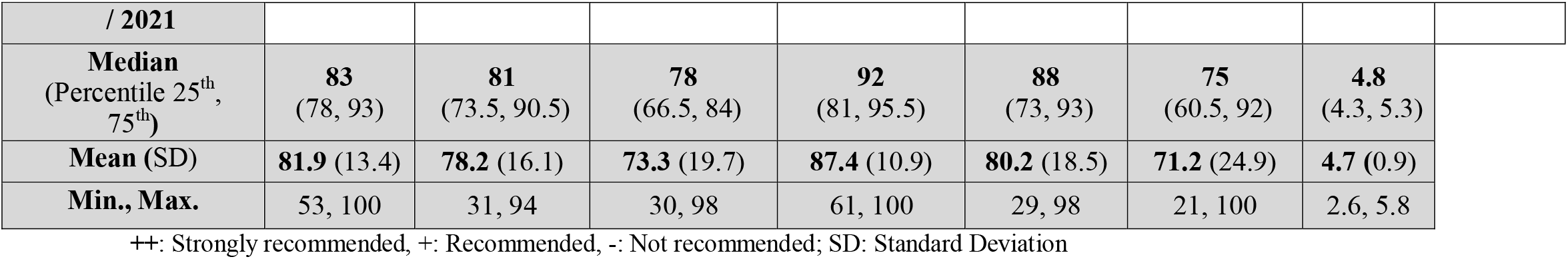
AGREE-II instrument’s six domain scores are expressed as a percentage.

**Figure 2** illustrates that the Editorial Independence domain of the “cardiovascular disease and Risk Management” guideline, developed in 2015 by the American Diabetic Association, was of the lowest score (**21**%) followed by the Rigour of Development (**30**%) of the same guideline, as well as the Applicability domain (**29**%) of “Guideline for Assessment of Cardiovascular Risk in Asymptomatic Adults” (ACCF/AHA 2010 Guideline). On the other hand, the majority of AGREE II domain scores that the included CVD screening guidelines achieved were above 60%, indicating that most of the included CVD screening guidelines were developed to a high-quality standard.

**Figure 2.**
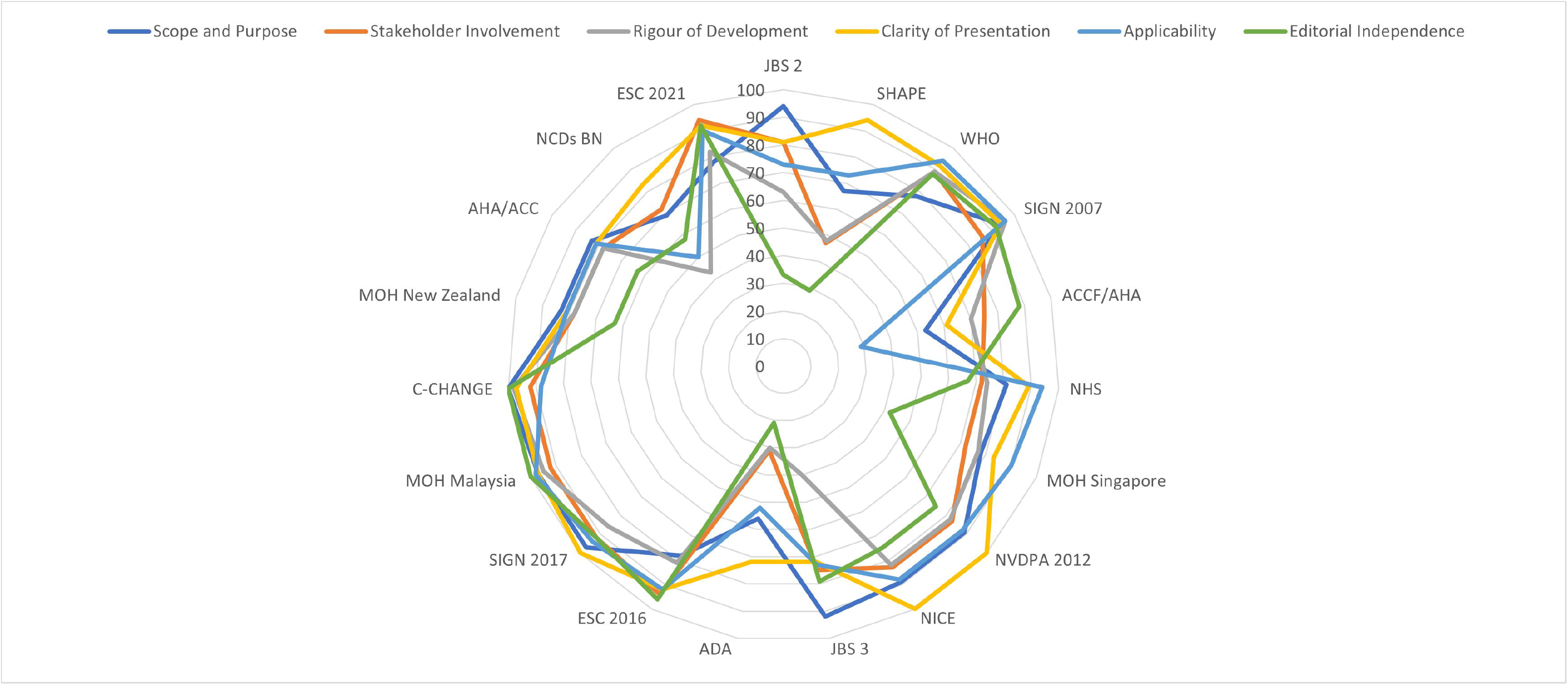

**Figure 3** shows that the highest quality guidelines in terms of AGREE II appraisal domains were “Primary & Secondary Prevention of Cardiovascular Disease” developed by the Ministry of Health Malaysia in 2017, followed by “Risk Estimation and the Prevention of cardiovascular disease” developed by the Scottish Intercollegiate Guidelines Network (SIGN) in 2007 and updated in 2017, “Canadian Cardiovascular Harmonized National Guidelines Endeavour (C-CHANGE) guideline for the prevention and management of cardiovascular disease in primary care” developed by The Canadian Cardiovascular Society in 2018 while the least quality guideline was “cardiovascular disease and Risk Management” guideline, developed in 2015 by American Diabetic Association.

19 out of the 20 guidelines were included in the analysis and evaluated using the AGREE II six domains. The recommendation system for the guidelines was then determined based on the domain scores, and 13 of the guidelines (**70%**) were classified as "strongly recommended" while only 6 (**30%**) were classified as just "recommended" as shown in **Table 2**.

**Figure 3.**
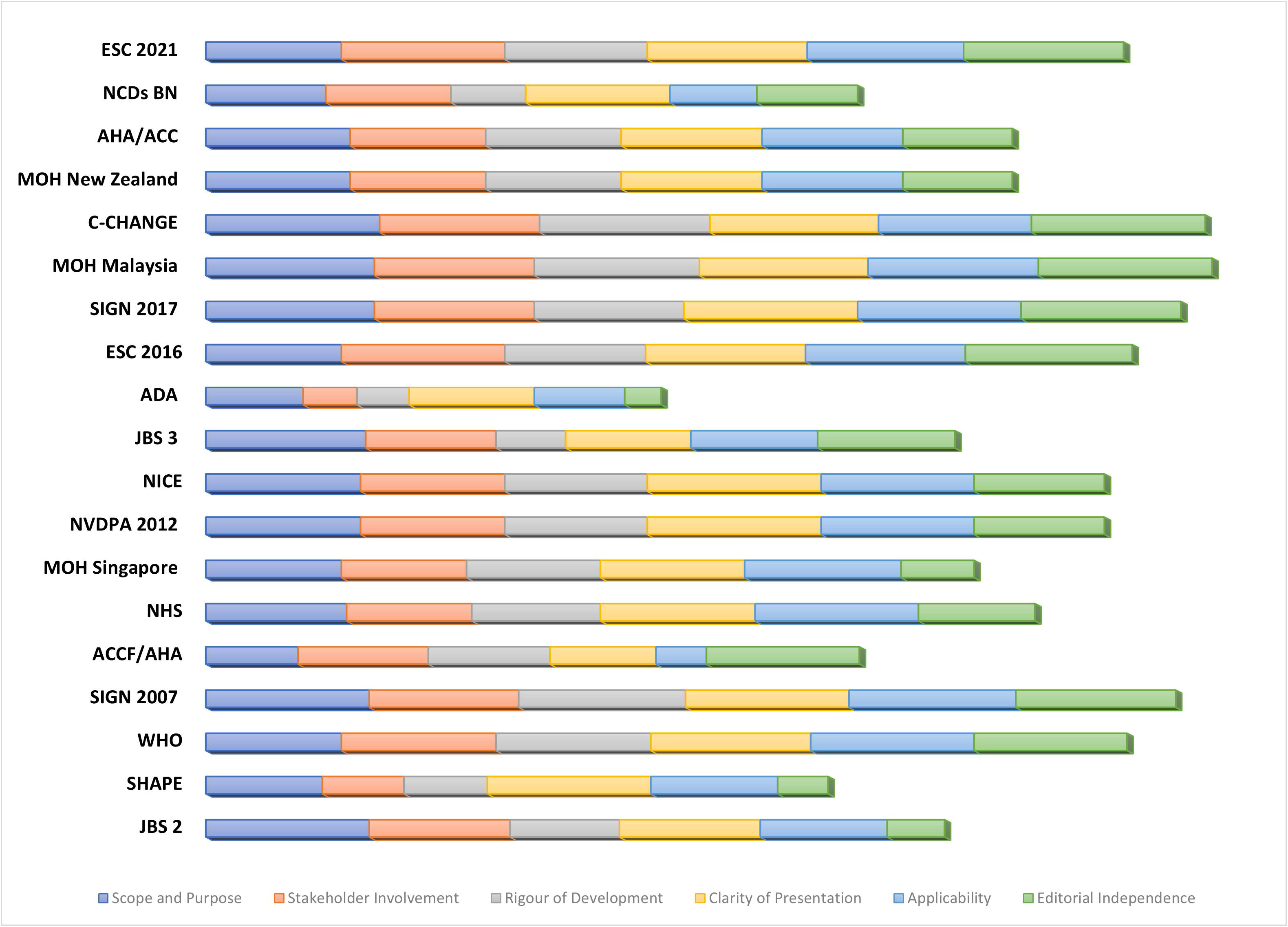

## DISCUSSION

### Summary of the Screening Guidelines CVD Prevention and Early Detection

To the best of our knowledge, the current scoping review represents the first evaluation of a substantial number of screening guidelines for the primary prevention and early diagnosis of cardiovascular disease. The CVD guidelines that are provided were developed by a widely diverse collection of international guideline developers.

Four of the 20 chosen CVD prevention guidelines were created in the United States. through a partnership between The American College of Cardiology Foundation and the American Heart Association or by independent efforts of the American Diabetes Association aiming to slow the progression of CVD risk in people with diabetes as well as the SHAPE (Screening for Heart Attack Prevention and Education) guideline which recommends noninvasive screening of all asymptomatic men and women aged 45 to 75 and 55 to 75, respectively (apart from those considered to be at extremely low risk) to identify and treat people who have subclinical atherosclerosis. In addition, the Canadian C-CHANGE guideline improves patient care in Canada by combining the recommendations of nine different guideline groups to assist physicians in creating a thorough CVD prevention and treatment strategy that is focused on the needs of the patients.

The chosen Eight European guidelines, including two continental ones, were created by the Task Force for Cardiovascular Disease Prevention in Clinical Practice, which included 12 medical societies and representatives of the European Society of Cardiology. The ESC chose the members of this Task Force, including participation from its pertinent ESC sub-speciality groups, to represent professionals involved in the medical management of CVD high-risk patients. Following ESC Clinical Practice Guidelines Committee (CPG) protocol, a group of qualified specialists reviewed all of the published data on cardiovascular disease prevention.

Additionally, the Scottish Intercollegiate Guidelines Network (SIGN) created two national Scottish guidelines to establish effective solutions for the reduction of CVD risk that directly affect the majority of the Scottish population. These guidelines mix a "high risk" and population approach. In addition, the United Kingdom created many guidelines, primarily for the Joint British Societies’ (JBS) recommendations for the prevention of cardiovascular disease in their second and third iterations. JBS2 prevention efforts often concentrate on patients with relatively low long-term (10-year) risks and certain pharmacologic therapy thresholds [18]. A unique aspect of JBS3 is the focus on the lifetime risk of CVD events, including a large pool of people in the population who have a lower 10-year risk of a CVD event but who nevertheless have a high lifetime event risk [25].

In Asia, there are three included guidelines developed by the Ministry of Health of Singapore, Malaysia, and Brunei Darussalam. Based on the best data that was available in each country at the time of development, these guidelines were created to serve as a national manual for optimal clinical practice in the prevention of cardiovascular disease. These Clinical Practice Guidelines are drawn up and usually updated by MOH national health screening related to cardiovascular disease and risk factors in each country to reflect the latest developments in the field and more current global targets set by the World Health Organization (WHO) [29].

Australian Guidelines for the management of Absolute cardiovascular disease risk are developed by the National Vascular Disease Prevention Alliance (NVDPA) which is an alliance of four leading and well-known Australian charities: Diabetes Australia, the National Heart Foundation of Australia, Kidney Health Australia and the National Stroke Foundation. In New Zealand, the Ministry of Health developed its CVD risk assessment and management guideline based on new five-year CVD risk prediction equations from New Zealand PREDICT study, to be known as the NZ Primary Prevention Equations [35].

Finally, the only global guideline from the included ones is the WHO guideline that was developed based on the total risk approach to the prevention of cardiovascular disease, elaborated in the World Health Report 2002 [36] which followed by preparations for the development of this guideline by the collaboration of International Society for Hypertension (ISH).

### Summary of AGREE II quality appraisal of CVD prevention guidelines

According to our literature search, this study is the first to evaluate this vast collection of screening guidelines for CVD prevention and early intervention. Our review made use of the AGREE II quality appraisal tool, which may be a helpful resource for several shareholders involved in decision-making, including **healthcare professionals** who prefer to do their own evaluation of a guideline before integrating its recommendations into their practice; **guideline developers** to follow a structured and rigorous development methodology and conduct a similar quality appraisal to ensure that their guidelines are sound or to evaluate guidelines from other groups for potential adaptation to their context; **policymakers** to assist them in deciding which guidelines could be recommended for use in practice or to inform policy decisions, and **educators** to help enhance critical appraisal skills amongst health professionals and to teach core competencies in guideline development and reporting [14].

According to our analysis, the majority of the included guidelines (70%) are highly advised for use by healthcare professionals as they performed well across the majority of AGREE II quality rating areas. The "cardiovascular disease and Risk Management" guideline, created in 2015 by the American Diabetic Association, was, however, the least trustworthy recommendation to be put into practice in the community. Even though it is primarily intended to reduce the risk of CVD in American adults with diabetes, the editorial independence domain’s quality is very lacking, so there is no assurance that funding body opinions won’t have an impact on the guideline’s content and that the developers’ group’s conflicts of interest have been disclosed and resolved.

On the other hand, due to their compliance with all requirements of the methodological quality appraisal outlined by the AGREE II instrument, Scottish guidelines developed by SIGN in 2007 and 2017, the Malaysian guideline developed by the Ministry of Health Malaysia in 2017 and the Canadian guideline developed in 2018 are the most reliable guidelines that health care professionals could use in their practice.

We observed that the best domain achieved by the included guidelines is the “Clarity of presentation” followed by the “Applicability” of the guidelines which entails that the CVD prevention guidelines’ developers pay close attention to the language, structure, and format of the guideline to be presented clearly to target users and the key recommendations and the options for managing CVD risk are provided clearly and unambiguously. Additionally, they ensure the applicability of the guidelines across health care providers by including enough information on the application facilitators and barriers; potential resource implications of applying the recommendations and clear monitoring criteria for the recommendations.

On the other hand, the least-achieved domains by the included guidelines are “Editorial Independence” and the “Rigour of development” which refers to the procedures used to gather and synthesize the evidence, the methods to formulate the recommendations and to update them [14]. Consistently with other overviews of clinical guidelines in many preventive medical fields, the content of the guidelines was influenced by the funding body’s viewpoint, and the heterogeneity of evidence and the strength of recommendation grading systems were lacking in this review similar to other clinical guideline overviews [37–40]. Another factor in the poor score for the included guidelines in the "Rigour of development" domain is that most of them did not specify a vigorous plan for updating their guidelines’ evidence based on reliable data from their own research, even though several of them noted that they should be reviewed every three or five years.

## CONCLUSION

In conclusion, we found numerous weaknesses in a number of guideline quality domains, exposing opportunities for quality improvement that future guideline developers should carefully explore and to be taken into consideration both now, when implementing the current guidelines, and in the future, when designing new ones.

We urge guideline developers to adopt AGREE-II international quality appraisal checklist to develop future sound CVD prevention guidelines. Guideline developers should ensure rigorous methodological processes and make recommendations that are formulated and disseminated in ways that ease understanding and application by the target users. When implementing or creating CVD guidelines, it is important to consider all of the AGREE-II domains that capture distinctive aspects of guideline quality, including "**Scope and Purpose**," "**Stakeholder Involvement**," "**Rigour of Development**," "**Clarity of Presentation**," and "**Editorial Independence**". Guidelines developers for future guidelines should put great concern on independence from funding bodies’ opinions besides updating their guideline’s evidence based on a vigorous plan of collecting reliable data from their own research.

## STRENGTHS AND LIMITATIONS

Our scoping review has several advantages, including the capacity to rate the methodological quality and the Rigour of development of the included guidelines using the AGREE II quality appraisal instrument. A thorough database search and reading of a sizable number of guidelines for CVD prevention and early detection were also conducted. A specified methodology and specific inclusion and exclusion criteria were also used in our search.

However, this review was limited by several potential limitations that should be taken into consideration. First, the quick review techniques and little biased assessment. Second, although the quality of the guideline was comprehensively assessed by a validated instrument (AGREE II tool), the AGREE II instrument only evaluates the reporting of the various components of the guidelines and not the content validity of their recommendations. A strong recommendation could be produced within a badly developed guideline, and vice versa, despite the fact that the quality of development across the entire guideline has a great potential to influence the quality of individual recommendations [41,42]. Third, the AGREE II tool only considered the information that was provided concerning the development of the guideline, and it did not completely rule out the potential that certain guidelines did not report detailed information in their guideline handbook, resulting in a lower AGREE II score.

## DECLARATIONS

### Ethics approval and consent to participate

As this review is based only on published studies, ethics approval and consent to participate are not applicable.

### Consent for publication

Not applicable.

### Availability of data and materials

All data analyzed during this study and supporting its findings are included in this published article and all studies included in this review are available in **Table 1**.

### Authors’ contributions

All authors contributed toward databases search, drafting, and critically revising the paper and agree to be accountable for all aspects of the work. The authors read and approved the final manuscript.

## Data Availability

All data produced in the present work are contained in the manuscript.

## Acknowledgements

This research is made possible through the generous support of the PAPRSB Institute of Health Sciences, Universiti Brunei Darussalam, Brunei Darussalam.

## ABBREVIATIONS

ACCF/AHA: The American College of Cardiology Foundation and the American Heart Association
ADA: American Diabetes Association
AGREE II: Appraisal of Guidelines for Research and Evaluation II
AHA/ACC: The American Heart Association and The American College of Cardiology
ASSIGN: ASsessing cardiovascular risk using SIGN guidelines to ASSIGN preventive treatment
CAD: coronary artery disease
C-CHANGE: Canadian Cardiovascular Harmonized National Guidelines Endeavour
CHD: Coronary heart disease.
CPG: Clinical practice guideline
CVD: Cardiovascular disease
CVDRA: CVD risk assessment
EAPC: European Association of Preventive Cardiology
ESC: European Society of Cardiology
GRADE: The Grading of Recommendations Assessment, Development and Evaluation
ISH: International Society of Hypertension
JBS: Joint British Societies’ Guidelines
Max.: Maximum
Min.: Minimum
MOH: Ministry of Health
NCDs: Noncommunicable Diseases
NCEP ATP: National Cholesterol Education Program’s Adult Treatment Panel
NHMRC: National Health and Medical Research Council
NHS: National Health Services
NICE: National Institute for Health and Care Excellence
NVDPA: The National Vascular Disease Prevention Alliance
PRISMA-ScR: Systematic reviews and Meta-Analyses extension for Scoping Reviews
SCORE: Systematic Coronary Risk Estimation
SCORE2-OP: Updated SCORE for old people
SHAPE: Screening for Heart Attack Prevention and Education
SIGN: Scottish Intercollegiate Guidelines Network
UK: United Kingdom
WHO: World Health Organization
WPR-A: Western Pacific Region A

