## Appendix 2 for "Quality Appraisal of Screening Guidelines for Cardiovascular disease Prevention: *a scoping review*"

### A critical group appraisal of:
### Primary Prevention of Cardiovascular Disease (NHS) / 2011
### using the AGREE II Instrument

Created with the AGREE II Online Guideline Appraisal Tool.

No endorsement of the content of this document by the AGREE Research Trust should be implied.

Co-ordinator: Mohammed Abd ElFattah Darwesh Badawy

Date: 30 November 2022

URL of this appraisal: http://www.agreetrust.org/group-appraisal/18172

Guideline URL:
http://www.nhsborders.scot.nhs.uk/media/197138/Primary-Prevention-of-Cardiovascular-Disease.pdf

| Domain 1 | Domain 2 | Domain 3 | Domain 4 | Domain 5 | Domain 6 | OA 1 | OA 2 |
|---|---|---|---|---|---|---|---|
| 81% | 72% | 74% | 89% | 94% | 67% | 58% | Yes - 0, Yes with modifications - 2, No - 0 |

| *Domain 1. Scope and Purpose* | | |
|---|---|---|
| | Appraiser 1 | Appraiser 3 |
| Item 1 | 6 | 6 |
| Item 2 | 7 | 7 |
| Item 3 | 5 | 4 |
| *Domain 2. Stakeholder Involvement* | | |
| | Appraiser 1 | Appraiser 3 |
| Item 4 | 6 | 4 |
| Item 5 | 5 | 5 |
| Item 6 | 6 | 6 |
| *Domain 3. Rigour of Development* | | |
| | Appraiser 1 | Appraiser 3 |
| Item 7 | 5 | 5 |
| Item 8 | 5 | 5 |
| Item 9 | 5 | 5 |
| Item 10 | 7 | 7 |
| Item 11 | 7 | 7 |
| Item 12 | 6 | 6 |
| Item 13 | 5 | 4 |
| Item 14 | 4 | 4 |
| *Domain 4. Clarity of Presentation* | | |
| | Appraiser 1 | Appraiser 3 |
| Item 15 | 6 | 6 |
| Item 16 | 6 | 6 |
| Item 17 | 7 | 7 |
| *Domain 5. Applicability* | | |
| | Appraiser 1 | Appraiser 3 |

| | | |
|---|---|---|
| Item 18 | 7 | 7 |
| Item 19 | 6 | 5 |
| Item 20 | 7 | 7 |
| Item 21 | 7 | 7 |
| *Domain 6. Editorial Independence* | | |
| | Appraiser 1 | Appraiser 3 |
| Item 22 | 4 | 4 |
| Item 23 | 6 | 6 |
| *Overall Assessment* | | |
| | Appraiser 1 | Appraiser 3 |
| OA1 | 5 | 4 |

Created online at www.agreetrust.org 30 November 2022

### A critical group appraisal of: Guidelines for the management of Absolute cardiovascular disease risk/ 2012 using the AGREE II Instrument

Created with the AGREE II Online Guideline Appraisal Tool.

No endorsement of the content of this document by the AGREE Research Trust should be implied.

Co-ordinator: Mohammed Abd ElFattah Darwesh Badawy

Date: 30 November 2022

URL of this appraisal: http://www.agreetrust.org/group-appraisal/18174

Guideline URL: https://www.heartfoundation.org.au/getmedia/4342a70f-4487-496e-bbb0-dae33a47fcb2/Absolute-CVD-Risk-Full-Guidelines_2.pdf

| Domain 1 | Domain 2 | Domain 3 | Domain 4 | Domain 5 | Domain 6 | OA 1 | OA 2 |
|---|---|---|---|---|---|---|---|
| 100% | 100% | 98% | 100% | 92% | 100% | 100% | Yes - 2, Yes with modifications - 0, No - 0 |

| *Domain 1. Scope and Purpose* | | |
|---|---|---|
| | Appraiser 1 | Appraiser 3 |
| Item 1 | 7 | 7 |
| Item 2 | 7 | 7 |
| Item 3 | 7 | 7 |
| *Domain 2. Stakeholder Involvement* | | |
| | Appraiser 1 | Appraiser 3 |
| Item 4 | 7 | 7 |
| Item 5 | 7 | 7 |
| Item 6 | 7 | 7 |
| *Domain 3. Rigour of Development* | | |
| | Appraiser 1 | Appraiser 3 |
| Item 7 | 7 | 7 |
| Item 8 | 7 | 7 |
| Item 9 | 6 | 6 |
| Item 10 | 7 | 7 |
| Item 11 | 7 | 7 |
| Item 12 | 7 | 7 |
| Item 13 | 7 | 7 |
| Item 14 | 7 | 7 |
| *Domain 4. Clarity of Presentation* | | |
| | Appraiser 1 | Appraiser 3 |
| Item 15 | 7 | 7 |
| Item 16 | 7 | 7 |
| Item 17 | 7 | 7 |
| *Domain 5. Applicability* | | |
| | Appraiser 1 | Appraiser 3 |

| | | |
|---|---|---|
| Item 18 | 7 | 7 |
| Item 19 | 7 | 7 |
| Item 20 | 6 | 6 |
| Item 21 | 6 | 6 |
| *Domain 6. Editorial Independence* | | |
| | Appraiser 1 | Appraiser 3 |
| Item 22 | 7 | 7 |
| Item 23 | 7 | 7 |
| *Overall Assessment* | | |
| | Appraiser 1 | Appraiser 3 |
| OA1 | 7 | 7 |

Created online at [www.agreetrust.org](www.agreetrust.org) 30 November 2022

### A critical group appraisal of: Cardiovascular disease: risk assessment and reduction, including li modificationpid / NICE 2014 using the AGREE II Instrument

Created with the AGREE II Online Guideline Appraisal Tool.

No endorsement of the content of this document by the AGREE Research Trust should be implied.

Co-ordinator: Mohammed Abd ElFattah Darwesh Badawy

Date: 30 November 2022

URL of this appraisal: http://www.agreetrust.org/group-appraisal/18175

Guideline URL:
https://www.nice.org.uk/guidance/cg181/resources/cardiovascular-disease-risk-assessment-and-reduction-including-lipid-modification-pdf-35109807660997

| Domain 1 | Domain 2 | Domain 3 | Domain 4 | Domain 5 | Domain 6 | OA 1 | OA 2 |
|---|---|---|---|---|---|---|---|
| 89% | 83% | 82% | 100% | 88% | 75% | 75% | Yes - 1, Yes with modifications - 1, No - 0 |

| *Domain 1. Scope and Purpose* | | |
|---|---|---|
| | Appraiser 1 | Appraiser 3 |
| Item 1 | 7 | 7 |
| Item 2 | 5 | 5 |
| Item 3 | 7 | 7 |
| *Domain 2. Stakeholder Involvement* | | |
| | Appraiser 1 | Appraiser 3 |
| Item 4 | 6 | 6 |
| Item 5 | 5 | 5 |
| Item 6 | 7 | 7 |
| *Domain 3. Rigour of Development* | | |
| | Appraiser 1 | Appraiser 3 |
| Item 7 | 5 | 5 |
| Item 8 | 5 | 5 |
| Item 9 | 5 | 5 |
| Item 10 | 7 | 5 |
| Item 11 | 7 | 7 |
| Item 12 | 6 | 5 |
| Item 13 | 7 | 7 |
| Item 14 | 7 | 7 |
| *Domain 4. Clarity of Presentation* | | |
| | Appraiser 1 | Appraiser 3 |
| Item 15 | 7 | 7 |
| Item 16 | 7 | 7 |
| Item 17 | 7 | 7 |
| *Domain 5. Applicability* | | |
| | Appraiser 1 | Appraiser 3 |

| | | |
|---|---|---|
| Item 18 | 6 | 6 |
| Item 19 | 7 | 7 |
| Item 20 | 7 | 7 |
| Item 21 | 5 | 5 |
| *Domain 6. Editorial Independence* | | |
| | Appraiser 1 | Appraiser 3 |
| Item 22 | 5 | 5 |
| Item 23 | 6 | 6 |
| *Overall Assessment* | | |
| | Appraiser 1 | Appraiser 3 |
| OA1 | 6 | 5 |

Created online at www.agreetrust.org 30 November 2022

### A critical group appraisal of: Screening for cardiovascular disease and risk factors – MOH clinical practice guidelines -Singapore using the AGREE II Instrument

Created with the AGREE II Online Guideline Appraisal Tool.

No endorsement of the content of this document by the AGREE Research Trust should be implied.

Co-ordinator: Mohammed Abd ElFattah Darwesh Badawy

Date: 12 July 2023

URL of this appraisal: http://www.agreetrust.org/group-appraisal/18173

Guideline URL:
https://www.moh.gov.sg/docs/librariesprovider4/guidelines/cpg_screening-for-cardiovascular-disease-mar-2011.pdf

| Domain 1 | Domain 2 | Domain 3 | Domain 4 | Domain 5 | Domain 6 | OA 1 | OA 2 |
|---|---|---|---|---|---|---|---|
| 78% | 72% | 77% | 83% | 90% | 42% | 50% | Yes - 0, Yes with modifications - 1, No - 0 |

| *Domain 1. Scope and Purpose* | | |
|---|---|---|
| | Appraiser 1 | Appraiser 4 |
| Item 1 | 5 | 6 |
| Item 2 | 6 | 6 |
| Item 3 | 6 | 5 |
| *Domain 2. Stakeholder Involvement* | | |
| | Appraiser 1 | Appraiser 4 |
| Item 4 | 5 | 5 |
| Item 5 | 5 | 5 |
| Item 6 | 6 | 6 |
| *Domain 3. Rigour of Development* | | |
| | Appraiser 1 | Appraiser 4 |
| Item 7 | 5 | 6 |
| Item 8 | 5 | 5 |
| Item 9 | 6 | 5 |
| Item 10 | 5 | 5 |
| Item 11 | 7 | 6 |
| Item 12 | 7 | 6 |
| Item 13 | 5 | 4 |
| Item 14 | 7 | 6 |
| *Domain 4. Clarity of Presentation* | | |
| | Appraiser 1 | Appraiser 4 |
| Item 15 | 6 | 6 |
| Item 16 | 6 | 5 |
| Item 17 | 7 | 6 |
| *Domain 5. Applicability* | | |
| | Appraiser 1 | Appraiser 4 |

| | | |
|---|---|---|
| Item 18 | 6 | 5 |
| Item 19 | 6 | 7 |
| Item 20 | 7 | 7 |
| Item 21 | 7 | 6 |
| *Domain 6. Editorial Independence* | | |
| | Appraiser 1 | Appraiser 4 |
| Item 22 | 4 | 3 |
| Item 23 | 4 | 3 |
| *Overall Assessment* | | |
| | Appraiser 1 | Appraiser 4 |
| OA1 | 5 | 3 |

Created online at www.agreetrust.org 12 July 2023

### A critical group appraisal of:
### JBS 3: Joint British Societies' Guidelines On Prevention of Cardiovascular Disease in Clinical Practice using the AGREE II Instrument

Created with the AGREE II Online Guideline Appraisal Tool.

No endorsement of the content of this document by the AGREE Research Trust should be implied.

Co-ordinator: Mohammed Abd ElFattah Darwesh Badawy

Date: 31 July 2023

URL of this appraisal: http://www.agreetrust.org/group-appraisal/18176

Guideline URL:
https://drive.google.com/file/d/1VkyIsVd2IuFkOHo5Q22pr3FIXA99Q0YJ/view?usp=sharing

| Domain 1 | Domain 2 | Domain 3 | Domain 4 | Domain 5 | Domain 6 | OA 1 | OA 2 |
|---|---|---|---|---|---|---|---|
| 92% | 75% | 40% | 72% | 73% | 79% | 67% | Yes - 1, Yes with modifications - 1, No - 0 |

| *Domain 1. Scope and Purpose* | | |
|---|---|---|
| | Appraiser 1 | Appraiser 4 |
| Item 1 | 7 | 6 |
| Item 2 | 6 | 7 |
| Item 3 | 7 | 6 |
| *Domain 2. Stakeholder Involvement* | | |
| | Appraiser 1 | Appraiser 4 |
| Item 4 | 6 | 5 |
| Item 5 | 5 | 6 |
| Item 6 | 6 | 5 |
| *Domain 3. Rigour of Development* | | |
| | Appraiser 1 | Appraiser 4 |
| Item 7 | 2 | 3 |
| Item 8 | 2 | 2 |
| Item 9 | 3 | 2 |
| Item 10 | 3 | 3 |
| Item 11 | 6 | 5 |
| Item 12 | 6 | 7 |
| Item 13 | 2 | 2 |
| Item 14 | 2 | 4 |
| *Domain 4. Clarity of Presentation* | | |
| | Appraiser 1 | Appraiser 4 |
| Item 15 | 7 | 1 |
| Item 16 | 6 | 6 |
| Item 17 | 6 | 6 |
| *Domain 5. Applicability* | | |
| | Appraiser 1 | Appraiser 4 |

| | | |
|---|---|---|
| Item 18 | 6 | 5 |
| Item 19 | 6 | 6 |
| Item 20 | 4 | 3 |
| Item 21 | 7 | 6 |
| *Domain 6. Editorial Independence* | | |
| | Appraiser 1 | Appraiser 4 |
| Item 22 | 6 | 6 |
| Item 23 | 6 | 5 |
| *Overall Assessment* | | |
| | Appraiser 1 | Appraiser 4 |
| OA1 | 6 | 4 |

Created online at www.agreetrust.org 31 July 2023

### A critical group appraisal of:
### Risk estimation and the prevention of cardiovascular disease / 2017
### using the AGREE II Instrument

Created with the AGREE II Online Guideline Appraisal Tool.

No endorsement of the content of this document by the AGREE Research Trust should be implied.

Co-ordinator: Mohammed Abd ElFattah Darwesh Badawy

Date: 31 July 2023

URL of this appraisal: http://www.agreetrust.org/group-appraisal/18306

Guideline URL: https://www.sign.ac.uk/media/1085/sign149.pdf

| Domain 1 | Domain 2 | Domain 3 | Domain 4 | Domain 5 | Domain 6 | OA 1 | OA 2 |
|---|---|---|---|---|---|---|---|
| 97% | 92% | 86% | 100% | 94% | 92% | 83% | Yes - 2, Yes with modifications - 0, No - 0 |

| *Domain 1. Scope and Purpose* | | |
|---|---|---|
| | Appraiser 1 | Appraiser 4 |
| Item 1 | 7 | 7 |
| Item 2 | 7 | 6 |
| Item 3 | 7 | 7 |
| *Domain 2. Stakeholder Involvement* | | |
| | Appraiser 1 | Appraiser 4 |
| Item 4 | 7 | 6 |
| Item 5 | 7 | 6 |
| Item 6 | 7 | 6 |
| *Domain 3. Rigour of Development* | | |
| | Appraiser 1 | Appraiser 4 |
| Item 7 | 7 | 7 |
| Item 8 | 6 | 5 |
| Item 9 | 7 | 6 |
| Item 10 | 7 | 5 |
| Item 11 | 7 | 6 |
| Item 12 | 7 | 5 |
| Item 13 | 7 | 6 |
| Item 14 | 6 | 5 |
| *Domain 4. Clarity of Presentation* | | |
| | Appraiser 1 | Appraiser 4 |
| Item 15 | 7 | 7 |
| Item 16 | 7 | 7 |
| Item 17 | 7 | 7 |
| *Domain 5. Applicability* | | |
| | Appraiser 1 | Appraiser 4 |

| | | |
|---|---|---|
| Item 18 | 7 | 6 |
| Item 19 | 7 | 7 |
| Item 20 | 7 | 6 |
| Item 21 | 7 | 6 |
| *Domain 6. Editorial Independence* | | |
| | Appraiser 1 | Appraiser 4 |
| Item 22 | 6 | 7 |
| Item 23 | 6 | 7 |
| *Overall Assessment* | | |
| | Appraiser 1 | Appraiser 4 |
| OA1 | 6 | 6 |

Created online at www.agreetrust.org 31 July 2023

### A critical group appraisal of: Primary & Secondary Prevention of Cardiovascular Disease / 2017 using the AGREE II Instrument

Created with the AGREE II Online Guideline Appraisal Tool.

No endorsement of the content of this document by the AGREE Research Trust should be implied.

Co-ordinator: Mohammed Abd ElFattah Darwesh Badawy

Date: 1 August 2023

URL of this appraisal: http://www.agreetrust.org/group-appraisal/18307

Guideline URL: https://www.moh.gov.my/moh/resources/Penerbitan/CPG/CARDIOVASCULAR/3.pdf

| Domain 1 | Domain 2 | Domain 3 | Domain 4 | Domain 5 | Domain 6 | OA 1 | OA 2 |
|---|---|---|---|---|---|---|---|
| 97% | 92% | 95% | 97% | 98% | 100% | 92% | Yes - 2, Yes with modifications - 0, No - 0 |

| *Domain 1. Scope and Purpose* | | |
|---|---|---|
| | Appraiser 1 | Appraiser 4 |
| Item 1 | 7 | 6 |
| Item 2 | 7 | 7 |
| Item 3 | 7 | 7 |
| *Domain 2. Stakeholder Involvement* | | |
| | Appraiser 1 | Appraiser 4 |
| Item 4 | 7 | 6 |
| Item 5 | 7 | 6 |
| Item 6 | 7 | 6 |
| *Domain 3. Rigour of Development* | | |
| | Appraiser 1 | Appraiser 4 |
| Item 7 | 7 | 7 |
| Item 8 | 7 | 7 |
| Item 9 | 7 | 6 |
| Item 10 | 6 | 7 |
| Item 11 | 7 | 7 |
| Item 12 | 7 | 6 |
| Item 13 | 7 | 6 |
| Item 14 | 7 | 6 |
| *Domain 4. Clarity of Presentation* | | |
| | Appraiser 1 | Appraiser 4 |
| Item 15 | 7 | 7 |
| Item 16 | 7 | 7 |
| Item 17 | 7 | 6 |
| *Domain 5. Applicability* | | |
| | Appraiser 1 | Appraiser 4 |

| | | |
|---|---|---|
| Item 18 | 7 | 6 |
| Item 19 | 7 | 7 |
| Item 20 | 7 | 7 |
| Item 21 | 7 | 7 |
| *Domain 6. Editorial Independence* | | |
| | Appraiser 1 | Appraiser 4 |
| Item 22 | 7 | 7 |
| Item 23 | 7 | 7 |
| *Overall Assessment* | | |
| | Appraiser 1 | Appraiser 4 |
| OA1 | 7 | 6 |

Created online at www.agreetrust.org 1 August 2023

### A critical group appraisal of: Canadian Cardiovascular Harmonized National Guidelines Endeavour (C-CHANGE) guideline for the prevention and management of cardiovascular disease in primary care / 2018 using the AGREE II Instrument

Created with the AGREE II Online Guideline Appraisal Tool.

No endorsement of the content of this document by the AGREE Research Trust should be implied.

Co-ordinator: Mohammed Abd ElFattah Darwesh Badawy

Date: 1 August 2023

URL of this appraisal: http://www.agreetrust.org/group-appraisal/18308

Guideline URL: https://www.cmaj.ca/content/190/40/E1192.long

| Domain 1 | Domain 2 | Domain 3 | Domain 4 | Domain 5 | Domain 6 | OA 1 | OA 2 |
|---|---|---|---|---|---|---|---|
| 100% | 92% | 98% | 97% | 88% | 100% | 100% | Yes - 2, Yes with modifications - 0, No - 0 |

| *Domain 1. Scope and Purpose* | | |
|---|---|---|
| | Appraiser 1 | Appraiser 4 |
| Item 1 | 7 | 7 |
| Item 2 | 7 | 7 |
| Item 3 | 7 | 7 |
| *Domain 2. Stakeholder Involvement* | | |
| | Appraiser 1 | Appraiser 4 |
| Item 4 | 7 | 7 |
| Item 5 | 6 | 7 |
| Item 6 | 6 | 6 |
| *Domain 3. Rigour of Development* | | |
| | Appraiser 1 | Appraiser 4 |
| Item 7 | 7 | 7 |
| Item 8 | 7 | 7 |
| Item 9 | 7 | 6 |
| Item 10 | 7 | 6 |
| Item 11 | 7 | 7 |
| Item 12 | 7 | 7 |
| Item 13 | 7 | 7 |
| Item 14 | 7 | 7 |
| *Domain 4. Clarity of Presentation* | | |
| | Appraiser 1 | Appraiser 4 |
| Item 15 | 7 | 7 |
| Item 16 | 6 | 7 |
| Item 17 | 7 | 7 |
| *Domain 5. Applicability* | | |
| | Appraiser 1 | Appraiser 4 |

| | | |
|---|---|---|
| Item 18 | 6 | 6 |
| Item 19 | 7 | 6 |
| Item 20 | 6 | 7 |
| Item 21 | 6 | 6 |
| *Domain 6. Editorial Independence* | | |
| | Appraiser 1 | Appraiser 4 |
| Item 22 | 7 | 7 |
| Item 23 | 7 | 7 |
| *Overall Assessment* | | |
| | Appraiser 1 | Appraiser 4 |
| OA1 | 7 | 7 |

Created online at www.agreetrust.org 1 August 2023

### A critical group appraisal of: ESC Guidelines on cardiovascular disease prevention in clinical practice / 2021 using the AGREE II Instrument

Created with the AGREE II Online Guideline Appraisal Tool.

No endorsement of the content of this document by the AGREE Research Trust should be implied.

Co-ordinator: Mohammed Abd ElFattah Darwesh Badawy

Date: 1 August 2023

URL of this appraisal: http://www.agreetrust.org/group-appraisal/18332

Guideline URL: https://academic.oup.com/eurheartj/article/42/34/3227/6358713?login=true

| Domain 1 | Domain 2 | Domain 3 | Domain 4 | Domain 5 | Domain 6 | OA 1 | OA 2 |
|---|---|---|---|---|---|---|---|
| 78% | 94% | 82% | 92% | 90% | 92% | 92% | Yes - 2, Yes with modifications - 0, No - 0 |

| *Domain 1. Scope and Purpose* | | |
|---|---|---|
| | Appraiser 1 | Appraiser 4 |
| Item 1 | 6 | 5 |
| Item 2 | 6 | 6 |
| Item 3 | 6 | 5 |
| *Domain 2. Stakeholder Involvement* | | |
| | Appraiser 1 | Appraiser 4 |
| Item 4 | 7 | 6 |
| Item 5 | 7 | 6 |
| Item 6 | 7 | 7 |
| *Domain 3. Rigour of Development* | | |
| | Appraiser 1 | Appraiser 4 |
| Item 7 | 5 | 5 |
| Item 8 | 5 | 6 |
| Item 9 | 7 | 6 |
| Item 10 | 5 | 5 |
| Item 11 | 7 | 7 |
| Item 12 | 7 | 6 |
| Item 13 | 7 | 7 |
| Item 14 | 5 | 5 |
| *Domain 4. Clarity of Presentation* | | |
| | Appraiser 1 | Appraiser 4 |
| Item 15 | 7 | 6 |
| Item 16 | 6 | 6 |
| Item 17 | 7 | 7 |
| *Domain 5. Applicability* | | |
| | Appraiser 1 | Appraiser 4 |

| | | |
|---|---|---|
| Item 18 | 7 | 6 |
| Item 19 | 6 | 6 |
| Item 20 | 6 | 6 |
| Item 21 | 7 | 7 |
| *Domain 6. Editorial Independence* | | |
| | Appraiser 1 | Appraiser 4 |
| Item 22 | 7 | 6 |
| Item 23 | 7 | 6 |
| *Overall Assessment* | | |
| | Appraiser 1 | Appraiser 4 |
| OA1 | 7 | 6 |

Created online at www.agreetrust.org 1 August 2023

AGREE II

### A critical group appraisal of: National Health Screening Guideline on Noncommunicable Diseases (NCDs) / 2019 using the AGREE II Instrument

Created with the AGREE II Online Guideline Appraisal Tool.

No endorsement of the content of this document by the AGREE Research Trust should be implied.

Co-ordinator: Mohammed Abd ElFattah Darwesh Badawy

Date: 1 August 2023

URL of this appraisal: http://www.agreetrust.org/group-appraisal/18333

Guideline URL: https://www.moh.gov.bn/Shared Documents/MOH_National Health Screening Guideline on NCDs_23 Jul 2020.pdf

| Domain 1 | Domain 2 | Domain 3 | Domain 4 | Domain 5 | Domain 6 | OA 1 | OA 2 |
|---|---|---|---|---|---|---|---|
| 69% | 72% | 43% | 83% | 50% | 58% | 58% | Yes - 0, Yes with modifications - 2, No - 0 |

| *Domain 1. Scope and Purpose* | | |
|---|---|---|
| | Appraiser 1 | Appraiser 4 |
| Item 1 | 5 | 4 |
| Item 2 | 5 | 5 |
| Item 3 | 6 | 6 |
| *Domain 2. Stakeholder Involvement* | | |
| | Appraiser 1 | Appraiser 4 |
| Item 4 | 6 | 6 |
| Item 5 | 3 | 3 |
| Item 6 | 7 | 7 |
| *Domain 3. Rigour of Development* | | |
| | Appraiser 1 | Appraiser 4 |
| Item 7 | 5 | 4 |
| Item 8 | 3 | 2 |
| Item 9 | 3 | 2 |
| Item 10 | 3 | 4 |
| Item 11 | 5 | 6 |
| Item 12 | 3 | 4 |
| Item 13 | 3 | 3 |
| Item 14 | 3 | 4 |
| *Domain 4. Clarity of Presentation* | | |
| | Appraiser 1 | Appraiser 4 |
| Item 15 | 7 | 7 |
| Item 16 | 5 | 4 |
| Item 17 | 7 | 6 |
| *Domain 5. Applicability* | | |
| | Appraiser 1 | Appraiser 4 |

| | | |
|---|---|---|
| Item 18 | 3 | 4 |
| Item 19 | 3 | 3 |
| Item 20 | 6 | 6 |
| Item 21 | 3 | 4 |
| *Domain 6. Editorial Independence* | | |
| | Appraiser 1 | Appraiser 4 |
| Item 22 | 3 | 6 |
| Item 23 | 5 | 4 |
| *Overall Assessment* | | |
| | Appraiser 1 | Appraiser 4 |
| OA1 | 5 | 4 |

Created online at www.agreetrust.org 1 August 2023

### A critical group appraisal of: AHA/ACC Guideline on the Primary Prevention of Cardiovascular Disease / 2019 using the AGREE II Instrument

Created with the AGREE II Online Guideline Appraisal Tool.

No endorsement of the content of this document by the AGREE Research Trust should be implied.

Co-ordinator: Mohammed Abd ElFattah Darwesh Badawy

Date: 1 August 2023

URL of this appraisal: http://www.agreetrust.org/group-appraisal/18334

Guideline URL: https://www.ahajournals.org/doi/epub/10.1161/CIR.0000000000000678

| Domain 1 | Domain 2 | Domain 3 | Domain 4 | Domain 5 | Domain 6 | OA 1 | OA 2 |
|---|---|---|---|---|---|---|---|
| 92% | 86% | 76% | 89% | 92% | 50% | 75% | Yes - 0, Yes with modifications - 1, No - 0 |

| *Domain 1. Scope and Purpose* | | |
|---|---|---|
| | Appraiser 1 | Appraiser 4 |
| Item 1 | 7 | 7 |
| Item 2 | 6 | 7 |
| Item 3 | 6 | 6 |
| *Domain 2. Stakeholder Involvement* | | |
| | Appraiser 1 | Appraiser 4 |
| Item 4 | 7 | 6 |
| Item 5 | 6 | 6 |
| Item 6 | 6 | 6 |
| *Domain 3. Rigour of Development* | | |
| | Appraiser 1 | Appraiser 4 |
| Item 7 | 7 | 6 |
| Item 8 | 6 | 5 |
| Item 9 | 7 | 6 |
| Item 10 | 3 | 3 |
| Item 11 | 6 | 6 |
| Item 12 | 7 | 7 |
| Item 13 | 7 | 6 |
| Item 14 | 3 | 4 |
| *Domain 4. Clarity of Presentation* | | |
| | Appraiser 1 | Appraiser 4 |
| Item 15 | 6 | 6 |
| Item 16 | 7 | 6 |
| Item 17 | 7 | 6 |
| *Domain 5. Applicability* | | |
| | Appraiser 1 | Appraiser 4 |

| | | |
|---|---|---|
| Item 18 | 7 | 7 |
| Item 19 | 6 | 6 |
| Item 20 | 7 | 7 |
| Item 21 | 6 | 6 |
| *Domain 6. Editorial Independence* | | |
| | Appraiser 1 | Appraiser 4 |
| Item 22 | 3 | 4 |
| Item 23 | 5 | 4 |
| *Overall Assessment* | | |
| | Appraiser 1 | Appraiser 4 |
| OA1 | 6 | 5 |

Created online at www.agreetrust.org 1 August 2023

### A critical group appraisal of: Cardiovascular Disease Risk Assessment and Management for Primary Care / 2018 using the AGREE II Instrument

Created with the AGREE II Online Guideline Appraisal Tool.

No endorsement of the content of this document by the AGREE Research Trust should be implied.

Co-ordinator: Mohammed Abd ElFattah Darwesh Badawy

Date: 1 August 2023

URL of this appraisal: http://www.agreetrust.org/group-appraisal/18335

Guideline URL:
https://www.health.govt.nz/system/files/documents/publications/cardiovascular-disease-risk-assessment-management-primary-care-feb18-v4_0.pdf

| Domain 1 | Domain 2 | Domain 3 | Domain 4 | Domain 5 | Domain 6 | OA 1 | OA 2 |
|---|---|---|---|---|---|---|---|
| 83% | 78% | 78% | 81% | 81% | 63% | 75% | Yes - 0, Yes with modifications - 1, No - 0 |

| *Domain 1. Scope and Purpose* | | |
|---|---|---|
| | Appraiser 1 | Appraiser 4 |
| Item 1 | 6 | 6 |
| Item 2 | 6 | 6 |
| Item 3 | 6 | 6 |
| *Domain 2. Stakeholder Involvement* | | |
| | Appraiser 1 | Appraiser 4 |
| Item 4 | 5 | 5 |
| Item 5 | 5 | 5 |
| Item 6 | 7 | 7 |
| *Domain 3. Rigour of Development* | | |
| | Appraiser 1 | Appraiser 4 |
| Item 7 | 7 | 7 |
| Item 8 | 7 | 6 |
| Item 9 | 5 | 6 |
| Item 10 | 5 | 5 |
| Item 11 | 7 | 7 |
| Item 12 | 4 | 4 |
| Item 13 | 5 | 4 |
| Item 14 | 6 | 6 |
| *Domain 4. Clarity of Presentation* | | |
| | Appraiser 1 | Appraiser 4 |
| Item 15 | 6 | 5 |
| Item 16 | 6 | 6 |
| Item 17 | 6 | 6 |
| *Domain 5. Applicability* | | |
| | Appraiser 1 | Appraiser 4 |

| | | |
|---|---|---|
| Item 18 | 6 | 5 |
| Item 19 | 5 | 6 |
| Item 20 | 6 | 6 |
| Item 21 | 7 | 6 |
| *Domain 6. Editorial Independence* | | |
| | Appraiser 1 | Appraiser 4 |
| Item 22 | 5 | 4 |
| Item 23 | 5 | 5 |
| *Overall Assessment* | | |
| | Appraiser 1 | Appraiser 4 |
| OA1 | 5 | 6 |

Created online at www.agreetrust.org 1 August 2023

### A critical group appraisal of: Guideline for Assessment of Cardiovascular Risk in Asymptomatic Adults (ACCF/AHA Guideline) using the AGREE II Instrument

Created with the AGREE II Online Guideline Appraisal Tool.

No endorsement of the content of this document by the AGREE Research Trust should be implied.

Co-ordinator: Mohammed Abd ElFattah Darwesh Badawy

Date: 1 August 2023

URL of this appraisal: http://www.agreetrust.org/group-appraisal/17970

Guideline URL: https://www.acc.org/~/media/clinical/pdf-files/approved-pdfs/2012/08/01/13/18/cv-risk-pocket-guide.pdf

| Domain 1 | Domain 2 | Domain 3 | Domain 4 | Domain 5 | Domain 6 | OA 1 | OA 2 |
|---|---|---|---|---|---|---|---|
| 53% | 75% | 70% | 61% | 29% | 88% | 50% | Yes - 0, Yes with modifications - 1, No - 0 |

| *Domain 1. Scope and Purpose* | | |
|---|---|---|
| | Appraiser 2 | Appraiser 4 |
| Item 1 | 5 | 4 |
| Item 2 | 3 | 3 |
| Item 3 | 6 | 4 |
| *Domain 2. Stakeholder Involvement* | | |
| | Appraiser 2 | Appraiser 4 |
| Item 4 | 6 | 5 |
| Item 5 | 5 | 4 |
| Item 6 | 7 | 6 |
| *Domain 3. Rigour of Development* | | |
| | Appraiser 2 | Appraiser 4 |
| Item 7 | 6 | 5 |
| Item 8 | 6 | 6 |
| Item 9 | 3 | 2 |
| Item 10 | 4 | 4 |
| Item 11 | 5 | 4 |
| Item 12 | 7 | 6 |
| Item 13 | 6 | 6 |
| Item 14 | 7 | 6 |
| *Domain 4. Clarity of Presentation* | | |
| | Appraiser 2 | Appraiser 4 |
| Item 15 | 7 | 6 |
| Item 16 | 3 | 2 |
| Item 17 | 5 | 5 |
| *Domain 5. Applicability* | | |
| | Appraiser 2 | Appraiser 4 |

| | | |
|---|---|---|
| Item 18 | 2 | 2 |
| Item 19 | 2 | 3 |
| Item 20 | 5 | 5 |
| Item 21 | 2 | 1 |
| *Domain 6. Editorial Independence* | | |
| | Appraiser 2 | Appraiser 4 |
| Item 22 | 7 | 5 |
| Item 23 | 7 | 6 |
| *Overall Assessment* | | |
| | Appraiser 2 | Appraiser 4 |
| OA1 | 5 | 3 |

Created online at www.agreetrust.org 1 August 2023

### A critical group appraisal of: Guidelines for assessment and management of cardiovascular risk using the AGREE II Instrument

Created with the AGREE II Online Guideline Appraisal Tool.

No endorsement of the content of this document by the AGREE Research Trust should be implied.

Co-ordinator: Mohammed Abd ElFattah Darwesh Badawy

Date: 1 August 2023

URL of this appraisal: http://www.agreetrust.org/group-appraisal/17968

Guideline URL: https://www.paho.org/en/documents/who-prevention-cardiovascular-disease-guidelines-assessment-and-management-0

| Domain 1 | Domain 2 | Domain 3 | Domain 4 | Domain 5 | Domain 6 | OA 1 | OA 2 |
|---|---|---|---|---|---|---|---|
| 78% | 89% | 89% | 92% | 94% | 88% | 100% | Yes - 1, Yes with modifications - 0, No - 0 |

| *Domain 1. Scope and Purpose* | | |
|---|---|---|
| | Appraiser 2 | Appraiser 4 |
| Item 1 | 7 | 6 |
| Item 2 | 6 | 5 |
| Item 3 | 5 | 5 |
| *Domain 2. Stakeholder Involvement* | | |
| | Appraiser 2 | Appraiser 4 |
| Item 4 | 6 | 6 |
| Item 5 | 6 | 7 |
| Item 6 | 6 | 7 |
| *Domain 3. Rigour of Development* | | |
| | Appraiser 2 | Appraiser 4 |
| Item 7 | 7 | 7 |
| Item 8 | 6 | 6 |
| Item 9 | 7 | 7 |
| Item 10 | 7 | 7 |
| Item 11 | 7 | 7 |
| Item 12 | 7 | 7 |
| Item 13 | 7 | 7 |
| Item 14 | 3 | 2 |
| *Domain 4. Clarity of Presentation* | | |
| | Appraiser 2 | Appraiser 4 |
| Item 15 | 6 | 5 |
| Item 16 | 7 | 7 |
| Item 17 | 7 | 7 |
| *Domain 5. Applicability* | | |
| | Appraiser 2 | Appraiser 4 |

| | | |
|---|---|---|
| Item 18 | 6 | 6 |
| Item 19 | 6 | 7 |
| Item 20 | 7 | 7 |
| Item 21 | 7 | 7 |
| *Domain 6. Editorial Independence* | | |
| | Appraiser 2 | Appraiser 4 |
| Item 22 | 6 | 6 |
| Item 23 | 6 | 7 |
| *Overall Assessment* | | |
| | Appraiser 2 | Appraiser 4 |
| OA1 | 7 | 7 |

Created online at www.agreetrust.org 1 August 2023

### A critical group appraisal of: SHAPE (Screening for Heart Attack Prevention and Education) Guideline using the AGREE II Instrument

Created with the AGREE II Online Guideline Appraisal Tool.

No endorsement of the content of this document by the AGREE Research Trust should be implied.

Co-ordinator: Mohammed Abd ElFattah Darwesh Badawy

Date: 1 August 2023

URL of this appraisal: http://www.agreetrust.org/group-appraisal/17967

Guideline URL: https://pubmed.ncbi.nlm.nih.gov/18340236/

| Domain 1 | Domain 2 | Domain 3 | Domain 4 | Domain 5 | Domain 6 | OA 1 | OA 2 |
|---|---|---|---|---|---|---|---|
| 67% | 47% | 48% | 94% | 73% | 29% | 50% | Yes - 0, Yes with modifications - 1, No - 0 |

| *Domain 1. Scope and Purpose* | | |
|---|---|---|
| | Appraiser 2 | Appraiser 4 |
| Item 1 | 5 | 5 |
| Item 2 | 4 | 3 |
| Item 3 | 7 | 6 |
| *Domain 2. Stakeholder Involvement* | | |
| | Appraiser 2 | Appraiser 4 |
| Item 4 | 5 | 5 |
| Item 5 | 3 | 3 |
| Item 6 | 3 | 4 |
| *Domain 3. Rigour of Development* | | |
| | Appraiser 2 | Appraiser 4 |
| Item 7 | 3 | 3 |
| Item 8 | 3 | 4 |
| Item 9 | 3 | 3 |
| Item 10 | 3 | 3 |
| Item 11 | 6 | 5 |
| Item 12 | 5 | 4 |
| Item 13 | 3 | 2 |
| Item 14 | 6 | 6 |
| *Domain 4. Clarity of Presentation* | | |
| | Appraiser 2 | Appraiser 4 |
| Item 15 | 7 | 6 |
| Item 16 | 7 | 6 |
| Item 17 | 7 | 7 |
| *Domain 5. Applicability* | | |
| | Appraiser 2 | Appraiser 4 |

| | | |
|---|---|---|
| Item 18 | 3 | 3 |
| Item 19 | 6 | 5 |
| Item 20 | 6 | 6 |
| Item 21 | 7 | 7 |
| *Domain 6. Editorial Independence* | | |
| | Appraiser 2 | Appraiser 4 |
| Item 22 | 3 | 3 |
| Item 23 | 3 | 2 |
| *Overall Assessment* | | |
| | Appraiser 2 | Appraiser 4 |
| OA1 | 5 | 3 |

Created online at www.agreetrust.org 1 August 2023

### A critical group appraisal of:
### JBS 2: Joint British Societies’ Guidelines On Prevention of Cardiovascular Disease in Clinical Practice using the AGREE II Instrument

Created with the AGREE II Online Guideline Appraisal Tool.

No endorsement of the content of this document by the AGREE Research Trust should be implied.

Co-ordinator: Mohammed Abd ElFattah Darwesh Badawy

Date: 1 August 2023

URL of this appraisal: http://www.agreetrust.org/group-appraisal/17966

Guideline URL: http://dx.doi.org/10.1136/hrt.2005.079988

| Domain 1 | Domain 2 | Domain 3 | Domain 4 | Domain 5 | Domain 6 | OA 1 | OA 2 |
|---|---|---|---|---|---|---|---|
| 94% | 81% | 63% | 81% | 73% | 33% | 75% | Yes - 1, Yes with modifications - 0, No - 0 |

| *Domain 1. Scope and Purpose* | | |
|---|---|---|
| | Appraiser 3 | Appraiser 5 |
| Item 1 | 7 | 7 |
| Item 2 | 6 | 7 |
| Item 3 | 7 | 6 |
| *Domain 2. Stakeholder Involvement* | | |
| | Appraiser 3 | Appraiser 5 |
| Item 4 | 7 | 6 |
| Item 5 | 5 | 6 |
| Item 6 | 5 | 6 |
| *Domain 3. Rigour of Development* | | |
| | Appraiser 3 | Appraiser 5 |
| Item 7 | 2 | 7 |
| Item 8 | 5 | 6 |
| Item 9 | 3 | 5 |
| Item 10 | 3 | 5 |
| Item 11 | 6 | 6 |
| Item 12 | 5 | 6 |
| Item 13 | 6 | 5 |
| Item 14 | 2 | 4 |
| *Domain 4. Clarity of Presentation* | | |
| | Appraiser 3 | Appraiser 5 |
| Item 15 | 6 | 6 |
| Item 16 | 7 | 5 |
| Item 17 | 5 | 6 |
| *Domain 5. Applicability* | | |
| | Appraiser 3 | Appraiser 5 |

| | | |
|---|---|---|
| Item 18 | 6 | 5 |
| Item 19 | 6 | 6 |
| Item 20 | 3 | 4 |
| Item 21 | 7 | 6 |
| *Domain 6. Editorial Independence* | | |
| | Appraiser 3 | Appraiser 5 |
| Item 22 | 3 | 3 |
| Item 23 | 3 | 3 |
| *Overall Assessment* | | |
| | Appraiser 3 | Appraiser 5 |
| OA1 | 6 | 5 |

Created online at www.agreetrust.org 1 August 2023

### A critical group appraisal of: Risk estimation and the prevention of cardiovascular disease SIGN 2007 using the AGREE II Instrument

Created with the AGREE II Online Guideline Appraisal Tool.

No endorsement of the content of this document by the AGREE Research Trust should be implied.

Co-ordinator: Mohammed Abd ElFattah Darwesh Badawy

Date: 1 August 2023

URL of this appraisal: http://www.agreetrust.org/group-appraisal/17969

Guideline URL:
https://drive.google.com/file/d/1B-k18hIS5rwwJjRY4FZZfLmnHBPW3BGw/view?usp=sharing

| Domain 1 | Domain 2 | Domain 3 | Domain 4 | Domain 5 | Domain 6 | OA 1 | OA 2 |
|---|---|---|---|---|---|---|---|
| 94% | 86% | 96% | 94% | 96% | 92% | 92% | Yes - 1, Yes with modifications - 0, No - 0 |

| *Domain 1. Scope and Purpose* | | |
|---|---|---|
| | Appraiser 2 | Appraiser 4 |
| Item 1 | 7 | 7 |
| Item 2 | 7 | 6 |
| Item 3 | 7 | 6 |
| *Domain 2. Stakeholder Involvement* | | |
| | Appraiser 2 | Appraiser 4 |
| Item 4 | 7 | 6 |
| Item 5 | 6 | 5 |
| Item 6 | 7 | 6 |
| *Domain 3. Rigour of Development* | | |
| | Appraiser 2 | Appraiser 4 |
| Item 7 | 7 | 7 |
| Item 8 | 7 | 7 |
| Item 9 | 7 | 6 |
| Item 10 | 7 | 7 |
| Item 11 | 7 | 6 |
| Item 12 | 7 | 6 |
| Item 13 | 7 | 6 |
| Item 14 | 7 | 7 |
| *Domain 4. Clarity of Presentation* | | |
| | Appraiser 2 | Appraiser 4 |
| Item 15 | 7 | 6 |
| Item 16 | 7 | 7 |
| Item 17 | 7 | 6 |
| *Domain 5. Applicability* | | |
| | Appraiser 2 | Appraiser 4 |

| | | |
|---|---|---|
| Item 18 | 7 | 7 |
| Item 19 | 7 | 7 |
| Item 20 | 7 | 6 |
| Item 21 | 7 | 6 |
| *Domain 6. Editorial Independence* | | |
| | Appraiser 2 | Appraiser 4 |
| Item 22 | 7 | 6 |
| Item 23 | 7 | 6 |
| *Overall Assessment* | | |
| | Appraiser 2 | Appraiser 4 |
| OA1 | 7 | 6 |

Created online at www.agreetrust.org 1 August 2023

### A critical group appraisal of:
### European Guidelines on cardiovascular disease prevention in clinical practice / 2016
### using the AGREE II Instrument

Created with the AGREE II Online Guideline Appraisal Tool.

No endorsement of the content of this document by the AGREE Research Trust should be implied.

Co-ordinator: Mohammed Abd ElFattah Darwesh Badawy

Date: 1 August 2023

URL of this appraisal: http://www.agreetrust.org/group-appraisal/18305

Guideline URL: https://academic.oup.com/eurheartj/article/37/29/2315/1748952

| Domain 1 | Domain 2 | Domain 3 | Domain 4 | Domain 5 | Domain 6 | OA 1 | OA 2 |
|---|---|---|---|---|---|---|---|
| 78% | 94% | 81% | 92% | 92% | 96% | 92% | Yes - 2, Yes with modifications - 0, No - 0 |

| *Domain 1. Scope and Purpose* | | |
|---|---|---|
| | Appraiser 1 | Appraiser 4 |
| Item 1 | 6 | 6 |
| Item 2 | 6 | 5 |
| Item 3 | 6 | 5 |
| *Domain 2. Stakeholder Involvement* | | |
| | Appraiser 1 | Appraiser 4 |
| Item 4 | 7 | 6 |
| Item 5 | 7 | 6 |
| Item 6 | 7 | 7 |
| *Domain 3. Rigour of Development* | | |
| | Appraiser 1 | Appraiser 4 |
| Item 7 | 5 | 6 |
| Item 8 | 5 | 6 |
| Item 9 | 7 | 6 |
| Item 10 | 5 | 4 |
| Item 11 | 7 | 7 |
| Item 12 | 7 | 6 |
| Item 13 | 7 | 7 |
| Item 14 | 5 | 4 |
| *Domain 4. Clarity of Presentation* | | |
| | Appraiser 1 | Appraiser 4 |
| Item 15 | 7 | 7 |
| Item 16 | 6 | 6 |
| Item 17 | 7 | 6 |
| *Domain 5. Applicability* | | |
| | Appraiser 1 | Appraiser 4 |

| | | |
|---|---|---|
| Item 18 | 7 | 6 |
| Item 19 | 6 | 6 |
| Item 20 | 6 | 7 |
| Item 21 | 7 | 7 |
| *Domain 6. Editorial Independence* | | |
| | Appraiser 1 | Appraiser 4 |
| Item 22 | 7 | 7 |
| Item 23 | 7 | 6 |
| *Overall Assessment* | | |
| | Appraiser 1 | Appraiser 4 |
| OA1 | 7 | 6 |

Created online at www.agreetrust.org 1 August 2023

### A critical group appraisal of: Cardiovascular Disease and Risk Management / 2015 using the AGREE II Instrument

Created with the AGREE II Online Guideline Appraisal Tool.

No endorsement of the content of this document by the AGREE Research Trust should be implied.

Co-ordinator: Mohammed Abd ElFattah Darwesh Badawy

Date: 1 August 2023

URL of this appraisal: http://www.agreetrust.org/group-appraisal/18304

Guideline URL: https://diabetesjournals.org/care/article/38/Supplement_1/S49/37273/8-Cardiovascular-Disease-and-Risk-Management

| Domain 1 | Domain 2 | Domain 3 | Domain 4 | Domain 5 | Domain 6 | OA 1 | OA 2 |
|---|---|---|---|---|---|---|---|
| 56% | 31% | 30% | 72% | 52% | 21% | 25% | Yes - 0, Yes with modifications - 1, No - 1 |

| *Domain 1. Scope and Purpose* | | |
|---|---|---|
| | Appraiser 1 | Appraiser 5 |
| Item 1 | 5 | 4 |
| Item 2 | 5 | 3 |
| Item 3 | 5 | 4 |
| *Domain 2. Stakeholder Involvement* | | |
| | Appraiser 1 | Appraiser 5 |
| Item 4 | 3 | 3 |
| Item 5 | 2 | 2 |
| Item 6 | 3 | 4 |
| *Domain 3. Rigour of Development* | | |
| | Appraiser 1 | Appraiser 5 |
| Item 7 | 2 | 1 |
| Item 8 | 2 | 2 |
| Item 9 | 2 | 1 |
| Item 10 | 2 | 3 |
| Item 11 | 6 | 5 |
| Item 12 | 6 | 6 |
| Item 13 | 2 | 1 |
| Item 14 | 2 | 2 |
| *Domain 4. Clarity of Presentation* | | |
| | Appraiser 1 | Appraiser 5 |
| Item 15 | 6 | 5 |
| Item 16 | 5 | 5 |
| Item 17 | 6 | 5 |
| *Domain 5. Applicability* | | |
| | Appraiser 1 | Appraiser 5 |

| | | |
|---|---|---|
| Item 18 | 5 | 6 |
| Item 19 | 5 | 5 |
| Item 20 | 2 | 2 |
| Item 21 | 6 | 2 |
| *Domain 6. Editorial Independence* | | |
| | Appraiser 1 | Appraiser 5 |
| Item 22 | 2 | 2 |
| Item 23 | 2 | 3 |
| *Overall Assessment* | | |
| | Appraiser 1 | Appraiser 5 |
| OA1 | 3 | 2 |

Created online at www.agreetrust.org 1 August 2023
