## Appendix 3 for "Quality Appraisal of Screening Guidelines for Cardiovascular disease Prevention: *a scoping review*"

### A critical appraisal of:
### JBS 2: Joint British Societies' Guidelines On Prevention of Cardiovascular Disease in Clinical Practice using the AGREE II Instrument

Created with the AGREE II Online Guideline Appraisal Tool.

No endorsement of the content of this document by the AGREE Research Trust should be implied.

Appraiser: Mohammed Abd ElFattah Darwesh Badawy

Date: 5 October 2022

URL of this appraisal: http://www.agreetrust.org/appraisal/96011

Guideline URL:

### Overall Assessment

Title: JBS 2: Joint British Societies' Guidelines On Prevention of Cardiovascular Disease in Clinical Practice

Overall quality of this guideline: 6/7

Guideline recommended for use? Yes.

| Domain | Total |
|---|---|
| 1. Scope and Purpose | 20 |
| 2. Stakeholder Involvement | 17 |
| 3. Rigour of Development | 32 |
| 4. Clarity of Presentation | 18 |
| 5. Applicability | 22 |
| 6. Editorial Independence | 6 |

---

### 1. Scope and Purpose

#### 1. The overall objective(s) of the guideline is (are) specifically described.

Rating: 7

The Objectives and priorities for cardiovascular disease prevention in clinical practice are described in detail in chapter II of this guideline with all the expected health benefits and defining the target population of this guideline.

#### 2. The health question(s) covered by the guideline is (are) specifically described.

Rating: 6

A detailed description of the health questions covered by the guideline is summarized in the summary and introduction chapters of this guideline. However, this was a little bit difficult to find within the text and the comparisons are not so evident.

#### 3. The population (patients, public, etc.) to whom the guideline is meant to apply is specifically described.

Rating: 7

A clear description of the population covered by a guideline is specifically provided with identifying groups of people who have equal priority for CVD prevention in clinical practice.

---

### 2. Stakeholder Involvement

#### 4. The guideline development group includes individuals from all relevant professional groups.

Rating: 7

Working Parties of the Joint British Societies' guidelines (JBS 2) working on cardiovascular disease prevention in clinical practice are listed in PREFACE with a concise and clear description of their roles.

#### 5. The views and preferences of the target population (patients, public, etc.) have been sought.

Rating: 5

Audit views have been added to each recommendation of this guideline. However, the methodological process of capturing the public's views and preferences is not clearly described.

#### 6. The target users of the guideline are clearly defined.

Rating: 5

The target users of this guideline are mentioned as the primary care practitioners has the main responsibilities for the identification and management of apparently healthy individuals at high total risk of developing CVD. However, it was not easy to find this content in the guideline.

---

### 3. Rigour of Development

#### 7. Systematic methods were used to search for evidence.

Rating: 2

Details of the strategy used to search for evidence is not well-provided in this guideline.

#### 8. The criteria for selecting the evidence are clearly described.

Rating: 5

Criteria for including/excluding evidence identified by the search are briefly provided under the Evaluation of scientific evidence section.

#### 9. The strengths and limitations of the body of evidence are clearly described.

Rating: 3

Statements highlighting the strengths and limitations of the body of evidence are not clearly described within the text of the guideline and are not easy to find.

##### 10. The methods for formulating the recommendations are clearly described.

Rating: 3

A description of the methods used to formulate the recommendations and how final decisions arrived are not clearly provided.

##### 11. The health benefits, side effects, and risks have been considered in formulating the recommendations.

Rating: 6

The health benefits, side effects, and risks have been well-considered in formulating the recommendations.

##### 12. There is an explicit link between the recommendations and the supporting evidence.

Rating: 5

The link between the recommendations and the evidence on which they are based is written, but it is not easy to extract from the text of the recommendation.

##### 13. The guideline has been externally reviewed by experts prior to its publication.

Rating: 6

External reviews include experts in the clinical area as well as some methodological experts and also auditing including the target population is presented in the preface and inside the guideline.

##### 14. A procedure for updating the guideline is provided.

Rating: 2

The history of joint body formulation is mentioned, and how the guideline is evolved but the procedure for updating the guideline is not provided.

---

#### 4. Clarity of Presentation

##### 15. The recommendations are specific and unambiguous.

Rating: 6

The recommendations provide a concrete and precise description of which option is appropriate in which situation and in what population group.

##### 16. The different options for management of the condition or health issue are clearly presented.

Rating: 7

The different options for management of the condition or health issue are clearly presented.

##### 17. Key recommendations are easily identifiable.

Rating: 5

Many recommendations are provided, and some tables and flowcharts are illustrated, but the readers can not find the most relevant recommendations easily.

---

#### 5. Applicability

##### 18. The guideline describes facilitators and barriers to its application.

Rating: 6

The guideline gives an explicit description of facilitators and barriers to the application of each recommendation.

##### 19. The guideline provides advice and/or tools on how the recommendations can be put into practice.

Rating: 6

The guideline provides advice and tools on how the recommendations can be implemented.

##### 20. The potential resource implications of applying the recommendations have been considered.

Rating: 3

There is no clear discussion in the guideline of the potential impact of the recommendations on resources.

##### 21. The guideline presents monitoring and/or auditing criteria.

Rating: 7

The guideline presents explicit monitoring and auditing criteria.

---

### 6. Editorial Independence

#### 22. The views of the funding body have not influenced the content of the guideline.

Rating: 3

There is no explicit statement that the views or interests of the funding body have not influenced the final recommendations.

#### 23. Competing interests of guideline development group members have been recorded and addressed.

Rating: 3

There is no explicit statement that all group members have declared whether they have any competing interests.

---

Created online at www.agreetrust.org 5 October 2022

### A critical appraisal of: SHAPE (Screening for Heart Attack Prevention and Education) Guideline using the AGREE II Instrument

Created with the AGREE II Online Guideline Appraisal Tool.

No endorsement of the content of this document by the AGREE Research Trust should be implied.

Appraiser: Mohammed Abd ElFattah Darwesh Badawy

Date: 8 October 2022

URL of this appraisal: http://www.agreetrust.org/appraisal/95318

Guideline URL:

---

#### Overall Assessment

Title: SHAPE (Screening for Heart Attack Prevention and Education) Guideline

Overall quality of this guideline: 5/7

Guideline recommended for use? Yes with modifications.

| Domain | Total |
|---|---|
| 1. Scope and Purpose | 16 |
| 2. Stakeholder Involvement | 11 |
| 3. Rigour of Development | 32 |
| 4. Clarity of Presentation | 21 |
| 5. Applicability | 22 |
| 6. Editorial Independence | 6 |

---

### 1. Scope and Purpose

#### 1. The overall objective(s) of the guideline is (are) specifically described.

Rating: 5

The objective of the SHAPE guideline is addressed as to detect and treat individuals with subclinical atherosclerosis; however, it was not easy to find in the text of the guideline.

#### 2. The health question(s) covered by the guideline is (are) specifically described.

Rating: 4

The health question is not clearly described in the body of the guideline.

#### 3. The population (patients, public, etc.) to whom the guideline is meant to apply is specifically described.

Rating: 7

The target population meant by the SHAPE guideline is explicitly described as all asymptomatic men 45 to 75 years old and asymptomatic women 55 to 75 years old (except those defined as very low risk).

---

### 2. Stakeholder Involvement

#### 4. The guideline development group includes individuals from all relevant professional groups.

Rating: 5

It\'s mentioned that The Screening for Heart Attack Prevention and Education (SHAPE)

Task Force is comprised of an international group of experts, but its international experts are not explicitly mentioned.

##### 5. The views and preferences of the target population (patients, public, etc.) have been sought.

Rating: 3

Information about the target population\'s experiences and expectations of health care are not well-informed in the guidelines.

##### 6. The target users of the guideline are clearly defined.

Rating: 3

The target users are not clearly defined in the guideline.

---

#### 3. Rigour of Development

##### 7. Systematic methods were used to search for evidence.

Rating: 3

Details of the strategy used to search for evidence are clearly described.

##### 8. The criteria for selecting the evidence are clearly described.

Rating: 3

Including/excluding criteria for selecting the evidence are not clearly described.

##### 9. The strengths and limitations of the body of evidence are clearly described.

Rating: 3

The strengths and limitations of the body of evidence are not clearly described

##### 10. The methods for formulating the recommendations are clearly described.

Rating: 3

The methods for formulating the recommendations are not clearly described.

##### 11. The health benefits, side effects, and risks have been considered in formulating the recommendations.

Rating: 6

The health benefits, side effects, and risks have been considered in formulating the recommendations.

##### 12. There is an explicit link between the recommendations and the supporting evidence.

Rating: 5

The link between the recommendations and the supporting evidence are found but not explicitly described.

##### 13. The guideline has been externally reviewed by experts prior to its publication.

Rating: 3

The external review of the SHAPE guideline is not mentioned.

##### 14. A procedure for updating the guideline is provided.

Rating: 6

A procedure for updating the guideline is provided, but the timescale of the update is not clear.

---

#### 4. Clarity of Presentation

##### 15. The recommendations are specific and unambiguous.

Rating: 7

The recommendations are specific and unambiguous.

##### 16. The different options for management of the condition or health issue are clearly presented.

Rating: 7

The different options for the management of the condition or health issue are clearly presented.

##### 17. Key recommendations are easily identifiable.

Rating: 7

Key recommendations are easily identifiable.

---

### 5. Applicability

##### 18. The guideline describes facilitators and barriers to its application.

Rating: 3

The SHAPE guideline lacks a description of facilitators and barriers to its application.

##### 19. The guideline provides advice and/or tools on how the recommendations can be put into practice.

Rating: 6

The guideline provides some tools on how the recommendations can be put into practice.

##### 20. The potential resource implications of applying the recommendations have been considered.

Rating: 6

In this era of limited healthcare resources, proof of cost-effectiveness of applying the recommendations is considered.

##### 21. The guideline presents monitoring and/or auditing criteria.

Rating: 7

The guideline presents explicit monitoring criteria.

---

### 6. Editorial Independence

##### 22. The views of the funding body have not influenced the content of the guideline.

Rating: 3

The influence of the views of the funding body is not mentioned in the guideline\'s content.

##### 23. Competing interests of guideline development group members have been recorded and addressed.

Rating: 3

The competing interests of guideline development group members have not been recorded and addressed.

---

Created online at www.agreetrust.org 8 October 2022

APPRAISAL OF GUIDELINES RESEARCH & EVALUATION

AGREE II

### A critical appraisal of: Guidelines for assessment and management of cardiovascular risk using the AGREE II Instrument

Created with the AGREE II Online Guideline Appraisal Tool.

No endorsement of the content of this document by the AGREE Research Trust should be implied.

Appraiser: Mohammed Abd ElFattah Darwesh Badawy

Date: 7 October 2022

URL of this appraisal: http://www.agreetrust.org/appraisal/95319

Guideline URL:

---

#### Overall Assessment

Title: Guidelines for assessment and management of cardiovascular risk

Overall quality of this guideline: 7/7

Guideline recommended for use? Yes.

| Domain | Total |
|---|---|
| 1. Scope and Purpose | 18 |
| 2. Stakeholder Involvement | 18 |
| 3. Rigour of Development | 51 |
| 4. Clarity of Presentation | 20 |
| 5. Applicability | 26 |
| 6. Editorial Independence | 12 |

---

### 1. Scope and Purpose

#### 1. The overall objective(s) of the guideline is (are) specifically described.

Rating: 7

The overall objective of this guideline is described thoroughly in the \"Background, scope and purpose of the guidelines\" section.

#### 2. The health question(s) covered by the guideline is (are) specifically described.

Rating: 6

The health question covered by the guideline is specifically described.

#### 3. The population (patients, public, etc.) to whom the guideline is meant to apply is specifically described.

Rating: 5

This guideline considers a framework, which can be adapted to suit different political, economic, social, cultural and medical circumstances.

---

### 2. Stakeholder Involvement

#### 4. The guideline development group includes individuals from all relevant professional groups.

Rating: 6

\"Development of the guidelines\" under the Introduction chapter describes The guideline development group including individuals from all relevant professional groups.

##### 5. The views and preferences of the target population (patients, public, etc.) have been sought.

Rating: 6

Information about the target population\\\'s experiences and expectations of health care is informed in the development of guidelines.

##### 6. The target users of the guideline are clearly defined.

Rating: 6

This guideline provides guidance to policy-makers and healthcare workers on how to target individuals at high risk of developing CVD at all levels of the health system and in different resource settings.

---

#### 3. Rigour of Development

##### 7. Systematic methods were used to search for evidence.

Rating: 7

All Details of the strategy used to search for evidence are provided in the section \"Development of the guideline\".

##### 8. The criteria for selecting the evidence are clearly described.

Rating: 6

The criteria for selecting the evidence are clearly described.

##### 9. The strengths and limitations of the body of evidence are clearly described.

Rating: 7

The strengths and limitations of the body of evidence are clearly described.

##### 10. The methods for formulating the recommendations are clearly described.

Rating: 7

Description of the methods used to formulate the recommendations and how final decisions were arrived at are clearly described.

##### 11. The health benefits, side effects, and risks have been considered in

**formulating the recommendations.**

Rating: 7

The health benefits, side effects, and risks have been considered in formulating the recommendations.

##### 12. There is an explicit link between the recommendations and the supporting evidence.

Rating: 7

There is an explicit link between the recommendations and the supporting evidence.

##### 13. The guideline has been externally reviewed by experts prior to its publication.

Rating: 7

The guideline is reviewed externally before its publication. Peer Reviewers have not been involved in the guideline development group.

##### 14. A procedure for updating the guideline is provided.

Rating: 3

A procedure for updating the guideline is not provided.

---

#### 4. Clarity of Presentation

##### 15. The recommendations are specific and unambiguous.

Rating: 6

The recommendations are specific and unambiguous.

##### 16. The different options for management of the condition or health issue are clearly presented.

Rating: 7

The different options for management of the condition or health issue are clearly presented.

##### 17. Key recommendations are easily identifiable.

Rating: 7

Key recommendations are easily identifiable.

#### 5. Applicability

##### 18. The guideline describes facilitators and barriers to its application.

Rating: 6

The guideline describes facilitators and barriers to its application.

##### 19. The guideline provides advice and/or tools on how the recommendations can be put into practice.

Rating: 6

The guideline provides advice tools on how the recommendations can be put into practice.

##### 20. The potential resource implications of applying the recommendations have been considered.

Rating: 7

Cost-effective preventive approaches to applying the recommendations have been considered.

##### 21. The guideline presents monitoring and/or auditing criteria.

Rating: 7

The guideline presents monitoring threshold criteria.

#### 6. Editorial Independence

##### 22. The views of the funding body have not influenced the content of the guideline.

Rating: 6

The views of the funding body have not influenced the content of the guideline

##### 23. Competing interests of guideline development group members have been recorded and addressed.

Rating: 6

The competing interests of guideline development group members have been recorded and addressed.

Created online at www.agreetrust.org 7 October 2022

### A critical appraisal of:
### Risk estimation and the prevention of cardiovascular disease
### using the AGREE II Instrument

Created with the AGREE II Online Guideline Appraisal Tool.

No endorsement of the content of this document by the AGREE Research Trust should be implied.

Appraiser: Mohammed Abd ElFattah Darwesh Badawy

Date: 7 October 2022

URL of this appraisal: http://www.agreetrust.org/appraisal/95320

Guideline URL:

---

#### Overall Assessment

Title: Risk estimation and the prevention of cardiovascular disease

Overall quality of this guideline: 7/7

Guideline recommended for use? Yes.

| Domain | Total |
|---|---|
| 1. Scope and Purpose | 21 |
| 2. Stakeholder Involvement | 20 |
| 3. Rigour of Development | 56 |
| 4. Clarity of Presentation | 21 |
| 5. Applicability | 28 |
| 6. Editorial Independence | 14 |

---

### 1. Scope and Purpose

#### 1. The overall objective(s) of the guideline is (are) specifically described.

Rating: 7

The overall objective of this guideline is described thoroughly in \"the need for a guideline\" section under the Introduction chapter.

#### 2. The health question(s) covered by the guideline is (are) specifically described.

Rating: 7

the guideline covers a detailed description of the health question in the introduction section.

#### 3. The population (patients, public, etc.) to whom the guideline is meant to apply is specifically described.

Rating: 7

A clear description of the population covered thoroughly by a guideline is provided.

---

### 2. Stakeholder Involvement

#### 4. The guideline development group includes individuals from all relevant professional groups.

Rating: 7

Chapter 14 \"Development of the guideline\" describes The guideline development group including individuals from all relevant professional groups.

##### 5. The views and preferences of the target population (patients, public, etc.) have been sought.

Rating: 6

Information about the target population\'s experiences and expectations of health care is informed in the development of guidelines.

##### 6. The target users of the guideline are clearly defined.

Rating: 7

The majority of the Scottish population as target users of the guideline are clearly defined.

---

#### 3. Rigour of Development

##### 7. Systematic methods were used to search for evidence.

Rating: 7

All Details of the strategy used to search for evidence are provided in the chapter of \"Development of the guideline\".

##### 8. The criteria for selecting the evidence are clearly described.

Rating: 7

The criteria for selecting the evidence are clearly described.

##### 9. The strengths and limitations of the body of evidence are clearly described.

Rating: 7

The grade of recommendation relates to the strength of the evidence, on which the recommendation is based, is clearly described.

##### 10. The methods for formulating the recommendations are clearly described.

Rating: 7

Description of the methods used to formulate the recommendations and how final decisions were arrived at are clearly described.

##### 11. The health benefits, side effects, and risks have been considered in formulating the recommendations.

Rating: 7

The health benefits, side effects, and risks have been considered in formulating the recommendations.

##### 12. There is an explicit link between the recommendations and the supporting evidence.

Rating: 7

There is an explicit link between the recommendations and the supporting evidence.

##### 13. The guideline has been externally reviewed by experts prior to its publication.

Rating: 7

A national open meeting is the main consultative phase of SIGN guideline development, at which the guideline development group present its draft recommendations for the first time.

##### 14. A procedure for updating the guideline is provided.

Rating: 7

A procedure for updating the guideline is obviously provided in the \"Introduction\".

---

#### 4. Clarity of Presentation

##### 15. The recommendations are specific and unambiguous.

Rating: 7

Recommendations provide concrete and precise descriptions of which option is appropriate in which situation and in what population group, as informed by the body of evidence.

##### 16. The different options for management of the condition or health issue are clearly presented.

Rating: 7

The different options for management of the condition or health issue are clearly presented.

##### 17. Key recommendations are easily identifiable.

Rating: 7

Key recommendations are easily identifiable.

---

### 5. Applicability

##### 18. The guideline describes facilitators and barriers to its application.

Rating: 7

The guideline describes the facilitators and barriers to its application.

##### 19. The guideline provides advice and/or tools on how the recommendations can be put into practice.

Rating: 7

The guideline provides tools on how the recommendations can be put into practice.

##### 20. The potential resource implications of applying the recommendations have been considered.

Rating: 7

The potential resource implications of applying the recommendations and economic costs have been considered.

##### 21. The guideline presents monitoring and/or auditing criteria.

Rating: 7

The guideline presents clear monitoring criteria for each recommendation.

---

### 6. Editorial Independence

##### 22. The views of the funding body have not influenced the content of the guideline.

Rating: 7

The views of the funding body have not influenced the content of the guideline.

##### 23. Competing interests of guideline development group members have been recorded and addressed.

Rating: 7

Competing interests of guideline development group members have been recorded and addressed.

---

Created online at www.agreetrust.org 7 October 2022

### A critical appraisal of: Guideline for Assessment of Cardiovascular Risk in Asymptomatic Adults (ACCF/AHA Guideline) using the AGREE II Instrument

Created with the AGREE II Online Guideline Appraisal Tool.

No endorsement of the content of this document by the AGREE Research Trust should be implied.

Appraiser: Mohammed Abd ElFattah Darwesh Badawy

Date: 4 October 2022

URL of this appraisal: [http://www.agreetrust.org/appraisal/95321](http://www.agreetrust.org/appraisal/95321)

Guideline URL:

---

### Overall Assessment

Title: Guideline for Assessment of Cardiovascular Risk in Asymptomatic Adults (ACCF/AHA Guideline)

Overall quality of this guideline: 5/7

Guideline recommended for use? Yes with modifications.

| Domain | Total |
|---|---|
| 1. Scope and Purpose | 14 |
| 2. Stakeholder Involvement | 18 |
| 3. Rigour of Development | 44 |
| 4. Clarity of Presentation | 15 |
| 5. Applicability | 11 |
| 6. Editorial Independence | 14 |

---

### 1. Scope and Purpose

#### 1. The overall objective(s) of the guideline is (are) specifically described.

Rating: 5

The overall objective of this guideline appears in the introduction section: \"The focus of this guideline is the initial assessment of the apparently healthy adult for risk of developing cardiovascular events associated with atherosclerotic vascular disease\". But the target population of this initial assessment is not clearly defined, and this item content is not straightforward to find within the guideline.

#### 2. The health question(s) covered by the guideline is (are) specifically described.

Rating: 3

The health question isn\'t clearly defined in the guideline. The assessment of CVD risk is understandable from the guidelines. Still, there is not enough information about the target population, comparative interventions or health settings where that assessment should be done.

#### 3. The population (patients, public, etc.) to whom the guideline is meant to apply is specifically described.

Rating: 6

The population is well defined at the start of the guidelines; This guideline should be applied to asymptomatic persons beginning at age 20, and the excluded populations with specific comorbidities are well described. However, the population information is not detailed enough so that the correct and eligible individuals would receive the action recommended.

---

### 2. Stakeholder Involvement

#### 4. The guideline development group includes individuals from all relevant professional groups.

Rating: 6

The guideline panel member list is included on the second page of the guideline with their qualification with each one\'s discipline, institution, location or role in the guideline is not well-illustrated.

#### 5. The views and preferences of the target population (patients, public, etc.) have been sought.

Rating: 5

Information about target population experiences and expectations of health care is not well-informed in the development of this guideline.

#### 6. The target users of the guideline are clearly defined.

Rating: 7

A clear description of the intended guideline target users is provided clearly in this guideline.

---

### 3. Rigour of Development

#### 7. Systematic methods were used to search for evidence.

Rating: 6

Details of the strategy used to search for evidence are well provided in this guideline.

#### 8. The criteria for selecting the evidence are clearly described.

Rating: 6

The criteria for applying the Classification of Recommendations according to the Level of Evidence are described. However, some detailed descriptions are missing in the inclusion or exclusion criteria for selecting the evidence.

##### 9. The strengths and limitations of the body of evidence are clearly described.

Rating: 3

The strengths and limitations of the body of evidence are stated briefly in table\'s caption but they are not clearly described

##### 10. The methods for formulating the recommendations are clearly described.

Rating: 4

The methods for formulating the recommendations are not clearly described in this guidelines

##### 11. The health benefits, side effects, and risks have been considered in formulating the recommendations.

Rating: 5

Health benefits and harms are considered in the recommendations according to the level of evidence given to each recommendation. However, supporting data and reporting of benefits/ harms are not clearly described.

##### 12. There is an explicit link between the recommendations and the supporting evidence.

Rating: 7

The recommendations and the supporting evidence are explicitly linked throughout the guideline.

##### 13. The guideline has been externally reviewed by experts prior to its publication.

Rating: 6

The external expert review is evident in this guideline.

##### 14. A procedure for updating the guideline is provided.

Rating: 7

An explicit procedure for updating the guideline is provided.

---

### 4. Clarity of Presentation

##### 15. The recommendations are specific and unambiguous.

Rating: 7

The recommendations are written in a specific, concrete and precise description.

##### 16. The different options for management of the condition or health issue are clearly presented.

Rating: 3

The different options for managing the CVD risk are not always considered.

##### 17. Key recommendations are easily identifiable.

Rating: 5

Some specific recommendations are grouped in one section, but the key recommendations are not clearly summarized.

---

#### 5. Applicability

##### 18. The guideline describes facilitators and barriers to its application.

Rating: 2

The existing facilitators and barriers that will impact the application of guideline recommendations are not clearly described.

##### 19. The guideline provides advice and/or tools on how the recommendations can be put into practice.

Rating: 2

This guideline does not provide advice and/or tools on how the recommendations can be implemented.

##### 20. The potential resource implications of applying the recommendations have been considered.

Rating: 5

There is some discussion about The potential additional resource implications of applying the recommendations.

##### 21. The guideline presents monitoring and/or auditing criteria.

Rating: 2

Monitoring and/or auditing criteria of each recommendation in this guideline are not clearly defined.

---

### 6. Editorial Independence

#### 22. The views of the funding body have not influenced the content of the guideline.

Rating: 7

it is prescribed that The Task Force makes every effort to avoid actual, potential, or perceived conflicts of interest that may arise as a result of industry relationships or personal interests among the writing committee.

#### 23. Competing interests of guideline development group members have been recorded and addressed.

Rating: 7

The Competing interests of guideline development group members have been recorded and addressed.

---

Created online at www.agreetrust.org 4 October 2022

### A critical appraisal of: Primary Prevention of Cardiovascular Disease (NHS) / 2011 using the AGREE II Instrument

Created with the AGREE II Online Guideline Appraisal Tool.

No endorsement of the content of this document by the AGREE Research Trust should be implied.

Appraiser: Mohammed Abd ElFattah Darwesh Badawy

Date: 23 October 2022

URL of this appraisal: http://www.agreetrust.org/appraisal/96211

Guideline URL:

---

#### Overall Assessment

Title: Primary Prevention of Cardiovascular Disease (NHS) / 2011

Overall quality of this guideline: 5/7

Guideline recommended for use? Yes with modifications.

| Domain | Total |
|---|---|
| 1. Scope and Purpose | 18 |
| 2. Stakeholder Involvement | 17 |
| 3. Rigour of Development | 44 |
| 4. Clarity of Presentation | 19 |
| 5. Applicability | 27 |
| 6. Editorial Independence | 10 |

---

### 1. Scope and Purpose

#### 1. The overall objective(s) of the guideline is (are) specifically described.

Rating: 6

The overall aim is described as updating the local policy for the primary prevention of cardiovascular disease (CVD) in high-risk groups.

#### 2. The health question(s) covered by the guideline is (are) specifically described.

Rating: 7

The health question is accurately described which is about the pros and cons of option 5, suggesting a ≥30% 10-year CVD risk for considering statins, as opposed to option 2, which suggests a ≥20% risk threshold.

#### 3. The population (patients, public, etc.) to whom the guideline is meant to apply is specifically described.

Rating: 5

The population to whom the guideline is meant to apply is described as the national population served by NHS. However, the detailed description is not described.

---

### 2. Stakeholder Involvement

#### 4. The guideline development group includes individuals from all relevant professional groups.

Rating: 6

The name of the author and the approval committee are provided in the guideline.

##### 5. The views and preferences of the target population (patients, public, etc.) have been sought.

Rating: 5

Evidence and trials are well-considered in this guideline, however, the views, preferences and expectations of the target population are not well-captured.

##### 6. The target users of the guideline are clearly defined.

Rating: 6

The target users of the guideline are clearly defined but hard to find to a clear section of text.

---

#### 3. Rigour of Development

##### 7. Systematic methods were used to search for evidence.

Rating: 5

Evidence and trials are well-considered in the guideline but the systematic methodology of the search is not described.

##### 8. The criteria for selecting the evidence are clearly described.

Rating: 5

The criteria for selecting the evidence are not so clearly described.

##### 9. The strengths and limitations of the body of evidence are clearly described.

Rating: 5

The strengths and limitations of the body of evidence are not clearly described.

##### 10. The methods for formulating the recommendations are clearly described.

Rating: 7

The methods for formulating the recommendations are clearly described in \"Consultation\" section

##### 11. The health benefits, side effects, and risks have been considered in

**formulating the recommendations.**

Rating: 7

The health benefits, side effects, and risks have been explicitly considered and described in formulating the recommendations.

##### 12. There is an explicit link between the recommendations and the supporting evidence.

Rating: 6

There is an explicit link between the recommendations and the supporting evidence.

##### 13. The guideline has been externally reviewed by experts prior to its publication.

Rating: 5

The external review is not explicitly considered in this guideline.

##### 14. A procedure for updating the guideline is provided.

Rating: 4

A procedure for updating the guideline is not provided.

---

#### 4. Clarity of Presentation

##### 15. The recommendations are specific and unambiguous.

Rating: 6

The recommendations are specific and unambiguous.

##### 16. The different options for management of the condition or health issue are clearly presented.

Rating: 6

The different options for management of the condition or health issue are clearly presented.

##### 17. Key recommendations are easily identifiable.

Rating: 7

Key recommendations are easily identifiable.

---

### 5. Applicability

##### 18. The guideline describes facilitators and barriers to its application.

Rating: 7

The guideline describes facilitators and barriers to its application.

##### 19. The guideline provides advice and/or tools on how the recommendations can be put into practice.

Rating: 6

The guideline provides clear advice on how the recommendations can be put into practice.

##### 20. The potential resource implications of applying the recommendations have been considered.

Rating: 7

The potential resource implications of applying the recommendations have been well-considered.

##### 21. The guideline presents monitoring and/or auditing criteria.

Rating: 7

The guideline presents clear monitoring and auditing criteria.

---

### 6. Editorial Independence

##### 22. The views of the funding body have not influenced the content of the guideline.

Rating: 4

The no explicit statement of no funding nor a statement that the funding body did not influence the content of the guideline.

##### 23. Competing interests of guideline development group members have been recorded and addressed.

Rating: 6

The competing interests of guideline development group members have been recorded and addressed.

---

### A critical appraisal of: Screening for cardiovascular disease and risk factors – MOH clinical practice guidelines -Singapore using the AGREE II Instrument

Created with the AGREE II Online Guideline Appraisal Tool.

No endorsement of the content of this document by the AGREE Research Trust should be implied.

Appraiser: Mohammed Abd ElFattah Darwesh Badawy

Date: 24 October 2022

URL of this appraisal: http://www.agreetrust.org/appraisal/96212

Guideline URL:

### Overall Assessment

Title: Screening for cardiovascular disease and risk factors – MOH clinical practice guidelines -Singapore

Overall quality of this guideline: 5/7

Guideline recommended for use? Yes with modifications.

| Domain | Total |
|---|---|
| 1. Scope and Purpose | 17 |
| 2. Stakeholder Involvement | 16 |
| 3. Rigour of Development | 47 |
| 4. Clarity of Presentation | 19 |
| 5. Applicability | 26 |
| 6. Editorial Independence | 8 |

---

### 1. Scope and Purpose

##### 1. The overall objective(s) of the guideline is (are) specifically described.

Rating: 5

The overall objective of the guideline is described, but it was not easy to find in the text.

##### 2. The health question(s) covered by the guideline is (are) specifically described.

Rating: 6

The health questions covered by the guideline are defined, but there was not enough information provided.

##### 3. The population (patients, public, etc.) to whom the guideline is meant to apply is specifically described.

Rating: 6

A clear description of the population covered by a guideline is provided.

---

### 2. Stakeholder Involvement

##### 4. The guideline development group includes individuals from all relevant professional groups.

Rating: 5

Working members for the development of this guideline are listed, but their roles are not clearly described.

##### 5. The views and preferences of the target population (patients, public, etc.) have been sought.

Rating: 5

The views and preferences of the target population have been sought, but there was no clear statement of the type of strategy used to capture these views.

##### 6. The target users of the guideline are clearly defined.

Rating: 6

The target users of the guideline are clearly defined.

---

#### 3. Rigour of Development

##### 7. Systematic methods were used to search for evidence.

Rating: 5

Systematic methods were used to search for evidence, but they are not clearly provided.

##### 8. The criteria for selecting the evidence are clearly described.

Rating: 5

The criteria for selecting the evidence are not clearly described.

##### 9. The strengths and limitations of the body of evidence are clearly described.

Rating: 6

The strengths and limitations of the body of evidence are clearly described.

##### 10. The methods for formulating the recommendations are clearly described.

Rating: 5

The methods for formulating the recommendations are not clearly described.

##### 11. The health benefits, side effects, and risks have been considered in formulating the recommendations.

Rating: 7

The health benefits, side effects, and risks have been considered in formulating the recommendations.

##### 12. There is an explicit link between the recommendations and the supporting evidence.

Rating: 7

There is an explicit link between the recommendations and the supporting evidence.

##### 13. The guideline has been externally reviewed by experts prior to its publication.

Rating: 5

The external review is not evident in this guideline.

##### 14. A procedure for updating the guideline is provided.

Rating: 7

A procedure for updating the guideline is provided as the working group advised for review every three years.

---

#### 4. Clarity of Presentation

##### 15. The recommendations are specific and unambiguous.

Rating: 6

The recommendations are specific and unambiguous.

##### 16. The different options for management of the condition or health issue are clearly presented.

Rating: 6

The different options for management of the condition or health issue are clearly presented.

##### 17. Key recommendations are easily identifiable.

Rating: 7

Key recommendations are easily identifiable in \"the Executive summary of the key recommendation.\"

### 5. Applicability

#### 18. The guideline describes facilitators and barriers to its application.

Rating: 6

The guideline describes the facilitators and barriers to its application.

#### 19. The guideline provides advice and/or tools on how the recommendations can be put into practice.

Rating: 6

The guideline provides clear advice on how the recommendations can be put into practice.

#### 20. The potential resource implications of applying the recommendations have been considered.

Rating: 7

The potential resource implications of applying the recommendations have been considered.

#### 21. The guideline presents monitoring and/or auditing criteria.

Rating: 7

The guideline presents the auditing criteria

---

### 6. Editorial Independence

#### 22. The views of the funding body have not influenced the content of the guideline.

Rating: 4

The views of the funding body are not clear in that guideline.

#### 23. Competing interests of guideline development group members have been recorded and addressed.

Rating: 4

The competing interests of guideline development group members have not been recorded and addressed.

Created online at www.agreetrust.org 24 October 2022

### A critical appraisal of:
### Guidelines for the management of Absolute cardiovascular disease risk/ 2012
### using the AGREE II Instrument

Created with the AGREE II Online Guideline Appraisal Tool.

No endorsement of the content of this document by the AGREE Research Trust should be implied.

Appraiser: Mohammed Abd ElFattah Darwesh Badawy

Date: 24 October 2022

URL of this appraisal: http://www.agreetrust.org/appraisal/96213

Guideline URL:

### Overall Assessment

Title: Guidelines for the management of Absolute cardiovascular disease risk/ 2012

Overall quality of this guideline: 7/7

Guideline recommended for use? Yes.

| Domain | Total |
|---|---|
| 1. Scope and Purpose | 21 |
| 2. Stakeholder Involvement | 21 |
| 3. Rigour of Development | 55 |
| 4. Clarity of Presentation | 21 |
| 5. Applicability | 26 |
| 6. Editorial Independence | 14 |

---

### 1. Scope and Purpose

#### 1. The overall objective(s) of the guideline is (are) specifically described.

Rating: 7

The overall objective of the guideline is specifically described and well-written in the \"scope\" section

#### 2. The health question(s) covered by the guideline is (are) specifically described.

Rating: 7

A detailed description of the health questions covered by the guideline is provided in a concise and clear presentation.

#### 3. The population (patients, public, etc.) to whom the guideline is meant to apply is specifically described.

Rating: 7

The population to whom the guideline is meant to apply is specifically and fully described in all detail.

---

### 2. Stakeholder Involvement

#### 4. The guideline development group includes individuals from all relevant professional groups.

Rating: 7

The guideline development group including individuals from all relevant professional groups are well-described in a separate section with details of their affiliations and a description of every member’s role.

#### 5. The views and preferences of the target population (patients, public, etc.) have been sought.

Rating: 7

The views and preferences of the target population have been sought and public consultation and comments are well-described in the guideline.

#### 6. The target users of the guideline are clearly defined.

Rating: 7

The target users of the guideline are clearly defined under the \"target audience\" section.

---

### 3. Rigour of Development

#### 7. Systematic methods were used to search for evidence.

Rating: 7

Systematic methods were used to search for evidence and details of the strategy used to search for evidence are provided including search terms used, sources consulted, and dates of the literature covered.

#### 8. The criteria for selecting the evidence are clearly described.

Rating: 7

Criteria for including/excluding evidence identified by the search are provided and clearly described.

#### 9. The strengths and limitations of the body of evidence are clearly described.

Rating: 6

The strengths and limitations of the body of evidence are clearly described.

##### 10. The methods for formulating the recommendations are clearly described.

Rating: 7

A description of the methods used to formulate the recommendations and how final decisions were arrived at are provided (modified Delphi technique).

##### 11. The health benefits, side effects, and risks have been considered in formulating the recommendations.

Rating: 7

The health benefits, side effects, and risks have been considered in formulating the recommendations.

##### 12. There is an explicit link between the recommendations and the supporting evidence.

Rating: 7

An explicit link between the recommendations and the supporting evidence is described in the guideline.

##### 13. The guideline has been externally reviewed by experts prior to its publication.

Rating: 7

Further refinement was undertaken after consultation with international guidelines groups in Scotland and New Zealand. Questions were then grouped under topics and circulated to experts for comment.

##### 14. A procedure for updating the guideline is provided.

Rating: 7

A procedure for updating the guideline is provided and clearly stated.

---

#### 4. Clarity of Presentation

##### 15. The recommendations are specific and unambiguous.

Rating: 7

Recommendations are provided with concrete and precise description of which option is appropriate in which situation and in what population group, as informed by the body of evidence.

##### 16. The different options for management of the condition or health issue are clearly presented.

Rating: 7

The different options for the management of the condition or health issue are clearly presented.

##### 17. Key recommendations are easily identifiable.

Rating: 7

Key recommendations are easily identifiable.

---

#### 5. Applicability

##### 18. The guideline describes facilitators and barriers to its application.

Rating: 7

The guideline describes the facilitators and barriers to its application.

##### 19. The guideline provides advice and/or tools on how the recommendations can be put into practice.

Rating: 7

The guideline provides clear advice and tools on how the recommendations can be put into practice.

##### 20. The potential resource implications of applying the recommendations have been considered.

Rating: 6

The potential resource implications of applying the recommendations have been considered.

##### 21. The guideline presents monitoring and/or auditing criteria.

Rating: 6

The guideline presents monitoring and auditing criteria.

---

#### 6. Editorial Independence

#### 22. The views of the funding body have not influenced the content of the guideline.

Rating: 7

The views of the funding body have not influenced the content of the guideline

#### 23. Competing interests of guideline development group members have been recorded and addressed.

Rating: 7

The competing interests of guideline development group members have been recorded and well addressed.

---

Created online at www.agreetrust.org 24 October 2022

### A critical appraisal of: Cardiovascular disease: risk assessment and reduction, including li modificationpid / NICE 2014 using the AGREE II Instrument

Created with the AGREE II Online Guideline Appraisal Tool.

No endorsement of the content of this document by the AGREE Research Trust should be implied.

Appraiser: Mohammed Abd ElFattah Darwesh Badawy

Date: 19 October 2022

URL of this appraisal: http://www.agreetrust.org/appraisal/96214

Guideline URL:

### Overall Assessment

Title: Cardiovascular disease: risk assessment and reduction, including li modificationpid / NICE 2014

Overall quality of this guideline: 6/7

Guideline recommended for use? Yes.

| Domain | Total |
|---|---|
| 1. Scope and Purpose | 19 |
| 2. Stakeholder Involvement | 18 |
| 3. Rigour of Development | 49 |
| 4. Clarity of Presentation | 21 |
| 5. Applicability | 25 |
| 6. Editorial Independence | 11 |

---

### 1. Scope and Purpose

#### 1. The overall objective(s) of the guideline is (are) specifically described.

Rating: 7

The clear aim of the guideline is described in the overview chapter as It aims to help healthcare professionals identify people who are at risk of cardiovascular problems including people with type 1 or type 2 diabetes, or chronic kidney disease.

#### 2. The health question(s) covered by the guideline is (are) specifically described.

Rating: 5

The health questions covered by the guideline are understandable, but they are not written in clear items.

#### 3. The population (patients, public, etc.) to whom the guideline is meant to apply is specifically described.

Rating: 7

A clear description of the population covered by a guideline is provided.

---

### 2. Stakeholder Involvement

#### 4. The guideline development group includes individuals from all relevant professional groups.

Rating: 6

The guideline development group includes individuals from all relevant professional groups and is listed in \"the details of the guideline committee\".

#### 5. The views and preferences of the target population (patients, public, etc.) have been sought.

Rating: 5

The views and preferences of the target population are considered, but the statement about the type of strategy used to capture publics' views and preferences is not well written.

#### 6. The target users of the guideline are clearly defined.

Rating: 7

The target users of the guideline are clearly defined

---

### 3. Rigour of Development

#### 7. Systematic methods were used to search for evidence.

Rating: 5

Systematic methods were used to search for evidence, but it is not well described in the guideline.

#### 8. The criteria for selecting the evidence are clearly described.

Rating: 5

The criteria for selecting the evidence are not clearly described.

#### 9. The strengths and limitations of the body of evidence are clearly described.

Rating: 5

he strengths and limitations of the body of evidence are not clearly described

#### 10. The methods for formulating the recommendations are clearly

**described.**

Rating: 7

The methods for updating the recommendations are clearly described in the Update information section and other sections.

##### 11. The health benefits, side effects, and risks have been considered in formulating the recommendations.

Rating: 7

The health benefits, side effects, and risks have been considered and well described in formulating the recommendations.

##### 12. There is an explicit link between the recommendations and the supporting evidence.

Rating: 6

There is a link between the recommendations and the supporting evidence, but need to be more explicit.

##### 13. The guideline has been externally reviewed by experts prior to its publication.

Rating: 7

Local commissioners and providers of healthcare have a responsibility to externally review and enable the guideline to be applied.

##### 14. A procedure for updating the guideline is provided.

Rating: 7

A procedure for updating the guideline is provided.

---

#### 4. Clarity of Presentation

##### 15. The recommendations are specific and unambiguous.

Rating: 7

The recommendations are specific and unambiguous.

##### 16. The different options for management of the condition or health issue are clearly presented.

Rating: 7

The different options for management of the condition or health issue are clearly presented.

##### 17. Key recommendations are easily identifiable.

Rating: 7

Key recommendations are easily identifiable under the chapter of \"Key priorities for implementation\".

---

#### 5. Applicability

##### 18. The guideline describes facilitators and barriers to its application.

Rating: 6

The guideline describes the facilitators and barriers to its application.

##### 19. The guideline provides advice and/or tools on how the recommendations can be put into practice.

Rating: 7

The guideline provides explicit tools on how the recommendations can be put into practice.

##### 20. The potential resource implications of applying the recommendations have been considered.

Rating: 7

The potential resource implications of applying the recommendations have been considered.

##### 21. The guideline presents monitoring and/or auditing criteria.

Rating: 5

The guideline does not present clear monitoring and/or auditing criteria with all recommendations.

---

#### 6. Editorial Independence

##### 22. The views of the funding body have not influenced the content of the guideline.

Rating: 5

There is no explicit statement that the views or interests of the funding body have not influenced the final recommendations.

#### 23. Competing interests of guideline development group members have been recorded and addressed.

Rating: 6

Competing interests of guideline development group members have been recorded and addressed.

---

Created online at www.agreetrust.org 19 October 2022

### A critical appraisal of:
### JBS 3: Joint British Societies’ Guidelines On Prevention of Cardiovascular Disease in Clinical Practice using the AGREE II Instrument

Created with the AGREE II Online Guideline Appraisal Tool.

No endorsement of the content of this document by the AGREE Research Trust should be implied.

Appraiser: Mohammed Abd ElFattah Darwesh Badawy

Date: 11 October 2022

URL of this appraisal: http://www.agreetrust.org/appraisal/96215

Guideline URL:

### Overall Assessment

Title: JBS 3: Joint British Societies' Guidelines On Prevention of Cardiovascular Disease in Clinical Practice

Overall quality of this guideline: 6/7

Guideline recommended for use? Yes.

| Domain | Total |
|---|---|
| 1. Scope and Purpose | 20 |
| 2. Stakeholder Involvement | 17 |
| 3. Rigour of Development | 26 |
| 4. Clarity of Presentation | 19 |
| 5. Applicability | 23 |
| 6. Editorial Independence | 12 |

---

### 1. Scope and Purpose

#### 1. The overall objective(s) of the guideline is (are) specifically described.

Rating: 7

The overall objective of the guideline is specifically described in Section 1 as the JBS3 guideline recommends a new approach to CVD prevention which builds on JBS2. It is not only based on 10-year absolute CVD risk but also CVD risk over an individual's lifetime.

#### 2. The health question(s) covered by the guideline is (are) specifically described.

Rating: 6

A detailed description of the health questions covered by the guideline is summarized in this guideline, and a comparison with JBS2 is evident. However, this was a little bit difficult to find within the text.

#### 3. The population (patients, public, etc.) to whom the guideline is meant to apply is specifically described.

Rating: 7

The population to whom the guideline is meant to apply is specifically described to include the population who are at low short term risk but at high lifetime risk.

---

### 2. Stakeholder Involvement

##### 4. The guideline development group includes individuals from all relevant professional groups.

Rating: 6

Working Parties of the Joint British Societies’ guidelines (JBS m) working on cardiovascular disease prevention in clinical practice are listed in Appendix 2.

##### 5. The views and preferences of the target population (patients, public, etc.) have been sought.

Rating: 5

Information about the target population\'s experiences and expectations of health care is not well-informed in the development of guidelines.

##### 6. The target users of the guideline are clearly defined.

Rating: 6

The target users of the guideline are clearly defined, primarily for healthcare professionals, to facilitate informed communication with individuals about their personal cardiovascular risk and the benefits of its early modification.

---

### 3. Rigour of Development

##### 7. Systematic methods were used to search for evidence.

Rating: 2

Details of the strategy used to search for evidence is not well-provided in this guideline.

##### 8. The criteria for selecting the evidence are clearly described.

Rating: 2

The criteria for selecting the evidence are not clearly described.

##### 9. The strengths and limitations of the body of evidence are clearly described.

Rating: 3

Statements highlighting the strengths and limitations of the body of evidence are not clearly described within the text of the guideline and are not easy to find.

##### 10. The methods for formulating the recommendations are clearly described.

Rating: 3

A description of the methods used to formulate the recommendations and how final decisions arrived are not clearly provided.

##### 11. The health benefits, side effects, and risks have been considered in formulating the recommendations.

Rating: 6

The health benefits, side effects, and risks have been well-considered in formulating the recommendations.

##### 12. There is an explicit link between the recommendations and the supporting evidence.

Rating: 6

The link between the recommendations and the evidence on which they are based is documented, but it is not easy to extract from the text of the recommendation.

##### 13. The guideline has been externally reviewed by experts prior to its publication.

Rating: 2

The external review is not evidently documented in the guideline.

##### 14. A procedure for updating the guideline is provided.

Rating: 2

A procedure for updating the guideline is not clearly provided.

---

#### 4. Clarity of Presentation

##### 15. The recommendations are specific and unambiguous.

Rating: 7

All recommendations are specific and unambiguous.

##### 16. The different options for management of the condition or health issue are clearly presented.

Rating: 6

The different options for the management of the condition or health issue are clearly presented.

##### 17. Key recommendations are easily identifiable.

Rating: 6

Many recommendations are provided, and some illustrations and examples are demonstrated.

---

#### 5. Applicability

##### 18. The guideline describes facilitators and barriers to its application.

Rating: 6

The guideline gives an explicit description of facilitators and barriers to the application of each recommendation.

##### 19. The guideline provides advice and/or tools on how the recommendations can be put into practice.

Rating: 6

The guideline provides advice and tools on how the recommendations can be implemented.

##### 20. The potential resource implications of applying the recommendations have been considered.

Rating: 4

Some cost implications are illustrated but, There is no clear discussion in the guideline of the potential impact of the recommendations on resources.

##### 21. The guideline presents monitoring and/or auditing criteria.

Rating: 7

The guideline presents explicit monitoring and auditing criteria.

---

#### 6. Editorial Independence

##### 22. The views of the funding body have not influenced the content of the guideline.

Rating: 6

There is an explicit statement that the views or interests of the funding body have not influenced the final recommendations.

#### 23. Competing interests of guideline development group members have been recorded and addressed.

Rating: 6

The competing interests of guideline development group members have been addressed.

---

Created online at www.agreetrust.org 11 October 2022

### A critical appraisal of: Cardiovascular Disease and Risk Management / 2015 using the AGREE II Instrument

Created with the AGREE II Online Guideline Appraisal Tool.

No endorsement of the content of this document by the AGREE Research Trust should be implied.

Appraiser: Mohammed Abd ElFattah Darwesh Badawy

Date: 26 October 2022

URL of this appraisal: http://www.agreetrust.org/appraisal/97106

Guideline URL:

---

#### Overall Assessment

Title: Cardiovascular Disease and Risk Management / 2015

Overall quality of this guideline: 3/7

Guideline recommended for use? Yes with modifications.

| Domain | Total |
|---|---|
| 1. Scope and Purpose | 15 |
| 2. Stakeholder Involvement | 8 |
| 3. Rigour of Development | 24 |
| 4. Clarity of Presentation | 17 |
| 5. Applicability | 18 |
| 6. Editorial Independence | 4 |

### 1. Scope and Purpose

#### 1. The overall objective(s) of the guideline is (are) specifically described.

Rating: 5

The overall objective of the guideline is described but not in detail.

#### 2. The health question(s) covered by the guideline is (are) specifically described.

Rating: 5

The health question covered by the guideline is described but not in detail.

#### 3. The population (patients, public, etc.) to whom the guideline is meant to apply is specifically described.

Rating: 5

A clear description of the population covered by a guideline is provided but not in a clear method.

### 2. Stakeholder Involvement

#### 4. The guideline development group includes individuals from all relevant professional groups.

Rating: 3

The guideline development group are not described in the text.

##### 5. The views and preferences of the target population (patients, public, etc.) have been sought.

Rating: 2

The views and preferences of the target population have not been sought.

##### 6. The target users of the guideline are clearly defined.

Rating: 3

The target users of the guideline are not clearly defined.

---

#### 3. Rigour of Development

##### 7. Systematic methods were used to search for evidence.

Rating: 2

Systematic methods used to search for evidence are not described.

##### 8. The criteria for selecting the evidence are clearly described.

Rating: 2

The criteria for selecting the evidence are not clearly described.

##### 9. The strengths and limitations of the body of evidence are clearly described.

Rating: 2

The strengths and limitations of the body of evidence are not clearly described.

##### 10. The methods for formulating the recommendations are clearly described.

Rating: 2

The methods for formulating the recommendations are not clearly described.

##### 11. The health benefits, side effects, and risks have been considered in formulating the recommendations.

Rating: 6

The health benefits, side effects, and risks have been considered in formulating the recommendations.

##### 12. There is an explicit link between the recommendations and the supporting evidence.

Rating: 6

There is an explicit link between the recommendations and the supporting evidence.

##### 13. The guideline has been externally reviewed by experts prior to its publication.

Rating: 2

The external review is not discussed in that guideline.

##### 14. A procedure for updating the guideline is provided.

Rating: 2

A procedure for updating the guideline is not provided.

---

#### 4. Clarity of Presentation

##### 15. The recommendations are specific and unambiguous.

Rating: 6

The recommendations are specific and unambiguous.

##### 16. The different options for management of the condition or health issue are clearly presented.

Rating: 5

The different options for the management of the condition or health issue are presented.

##### 17. Key recommendations are easily identifiable.

Rating: 6

Key recommendations are easily identifiable.

---

#### 5. Applicability

##### 18. The guideline describes facilitators and barriers to its application.

Rating: 5

The guideline describes the facilitators and barriers to its application but not clearly.

##### 19. The guideline provides advice and/or tools on how the recommendations can be put into practice.

Rating: 5

The guideline provides some advice on how the recommendations can be put into practice.

##### 20. The potential resource implications of applying the recommendations have been considered.

Rating: 2

The potential resource implications of applying the recommendations have not been considered.

##### 21. The guideline presents monitoring and/or auditing criteria.

Rating: 6

The guideline presents monitoring criteria.

---

#### 6. Editorial Independence

##### 22. The views of the funding body have not influenced the content of the guideline.

Rating: 2

The views of the funding body are not presented in the guideline.

##### 23. Competing interests of guideline development group members have been recorded and addressed.

Rating: 2

The competing interests of guideline development group members have not been recorded.

---

Created online at www.agreetrust.org 26 October 2022

### A critical appraisal of: European Guidelines on cardiovascular disease prevention in clinical practice / 2016 using the AGREE II Instrument

Created with the AGREE II Online Guideline Appraisal Tool.

No endorsement of the content of this document by the AGREE Research Trust should be implied.

Appraiser: Mohammed Abd ElFattah Darwesh Badawy

Date: 28 October 2022

URL of this appraisal: http://www.agreetrust.org/appraisal/97107

Guideline URL:

### Overall Assessment

Title: European Guidelines on cardiovascular disease prevention in clinical practice / 2016

Overall quality of this guideline: 7/7

Guideline recommended for use? Yes.

| Domain | Total |
|---|---|
| 1. Scope and Purpose | 18 |
| 2. Stakeholder Involvement | 21 |
| 3. Rigour of Development | 48 |
| 4. Clarity of Presentation | 20 |
| 5. Applicability | 26 |
| 6. Editorial Independence | 14 |

---

### 1. Scope and Purpose

#### 1. The overall objective(s) of the guideline is (are) specifically described.

Rating: 6

The overall objective of the guideline is specifically described but not easy to find in the text.

#### 2. The health question(s) covered by the guideline is (are) specifically described.

Rating: 6

The health question covered by the guideline is specifically described but not easy to find in the text.

#### 3. The population (patients, public, etc.) to whom the guideline is meant to apply is specifically described.

Rating: 6

The population to whom the guideline is meant to apply is specifically described.

---

### 2. Stakeholder Involvement

##### 4. The guideline development group includes individuals from all relevant professional groups.

Rating: 7

The guideline development group includes individuals from all relevant professional groups.

##### 5. The views and preferences of the target population (patients, public, etc.) have been sought.

Rating: 7

The views and preferences of the target population have been sought.

##### 6. The target users of the guideline are clearly defined.

Rating: 7

The target users of the guideline are clearly defined.

---

#### 3. Rigour of Development

##### 7. Systematic methods were used to search for evidence.

Rating: 5

Details of the strategy used to search for evidence are not clearly provided.

##### 8. The criteria for selecting the evidence are clearly described.

Rating: 5

The criteria for selecting the evidence are not clearly described.

##### 9. The strengths and limitations of the body of evidence are clearly described.

Rating: 7

The strengths and limitations of the body of evidence are clearly described.

##### 10. The methods for formulating the recommendations are clearly described.

Rating: 5

The methods for formulating the recommendations are not clearly described.

##### 11. The health benefits, side effects, and risks have been considered in formulating the recommendations.

Rating: 7

The health benefits, side effects, and risks have been considered in formulating the recommendations.

##### 12. There is an explicit link between the recommendations and the supporting evidence.

Rating: 7

There is an explicit link between the recommendations and the supporting evidence.

##### 13. The guideline has been externally reviewed by experts prior to its publication.

Rating: 7

The guideline has been externally reviewed by experts prior to its publication.

##### 14. A procedure for updating the guideline is provided.

Rating: 5

A procedure for updating the guideline is not clearly provided.

---

#### 4. Clarity of Presentation

##### 15. The recommendations are specific and unambiguous.

Rating: 7

The recommendations are specific and unambiguous.

##### 16. The different options for management of the condition or health issue are clearly presented.

Rating: 6

The different options for management of the condition or health issue are clearly presented.

##### 17. Key recommendations are easily identifiable.

Rating: 7

Key recommendations are easily identifiable.

### 5. Applicability

#### 18. The guideline describes facilitators and barriers to its application.

Rating: 7

The guideline describes the facilitators and barriers to its application.

#### 19. The guideline provides advice and/or tools on how the recommendations can be put into practice.

Rating: 6

The guideline provides advice on how the recommendations can be put into practice.

#### 20. The potential resource implications of applying the recommendations have been considered.

Rating: 6

The potential resource implications of applying the recommendations have been considered.

#### 21. The guideline presents monitoring and/or auditing criteria.

Rating: 7

The guideline presents monitoring and auditing criteria.

### 6. Editorial Independence

#### 22. The views of the funding body have not influenced the content of the guideline.

Rating: 7

The views of the funding body have not influenced the content of the guideline.

#### 23. Competing interests of guideline development group members have been recorded and addressed.

Rating: 7

Competing interests of guideline development group members have been recorded and addressed.

Created online at www.agreetrust.org 28 October 2022

### A critical appraisal of:
### Risk estimation and the prevention of cardiovascular disease / 2017
### using the AGREE II Instrument

Created with the AGREE II Online Guideline Appraisal Tool.

No endorsement of the content of this document by the AGREE Research Trust should be implied.

Appraiser: Mohammed Abd ElFattah Darwesh Badawy

Date: 26 October 2022

URL of this appraisal: http://www.agreetrust.org/appraisal/97114

Guideline URL:

---

#### Overall Assessment

Title: Risk estimation and the prevention of cardiovascular disease / 2017

Overall quality of this guideline: 6/7

Guideline recommended for use? Yes.

| Domain | Total |
|---|---|
| 1. Scope and Purpose | 21 |
| 2. Stakeholder Involvement | 21 |
| 3. Rigour of Development | 54 |
| 4. Clarity of Presentation | 21 |
| 5. Applicability | 28 |
| 6. Editorial Independence | 12 |

---

### 1. Scope and Purpose

#### 1. The overall objective(s) of the guideline is (are) specifically described.

Rating: 7

The overall objective of this guideline is described thoroughly in the \"REMIT of the guideline\" section under the Introduction chapter.

#### 2. The health question(s) covered by the guideline is (are) specifically described.

Rating: 7

The guideline covers a detailed description of the health question in the introduction section in the \"NEED for the guideline\" section.

#### 3. The population (patients, public, etc.) to whom the guideline is meant to apply is specifically described.

Rating: 7

A clear description of the population covered thoroughly by a guideline is provided.

---

### 2. Stakeholder Involvement

#### 4. The guideline development group includes individuals from all relevant professional groups.

Rating: 7

Chapter 16 \"Development of the guideline\" refers to the professionals who were involved at some stage of the development process. This includes members of the steering group, the research team involved in selecting and reviewing the evidence, and individuals involved in formulating the final recommendations.

##### 5. The views and preferences of the target population (patients, public, etc.) have been sought.

Rating: 7

The views and preferences of the target population are well-informed through the guidelines.

##### 6. The target users of the guideline are clearly defined.

Rating: 7

The target users of the guideline are clearly defined under the \"TARGET USERS OF THE GUIDELINE\" section.

---

#### 3. Rigour of Development

##### 7. Systematic methods were used to search for evidence.

Rating: 7

All Details of the strategy used to search for evidence are provided in chapter 15 \"SYSTEMATIC LITERATURE REVIEW.\"

##### 8. The criteria for selecting the evidence are clearly described.

Rating: 6

The criteria for selecting the evidence are described

##### 9. The strengths and limitations of the body of evidence are clearly described.

Rating: 7

The grade of recommendation relates to the strength of the evidence, on which the recommendation is based, is clearly described.

##### 10. The methods for formulating the recommendations are clearly described.

Rating: 7

Description of the methods used to formulate the recommendations and how final decisions were arrived at are clearly described.

##### 11. The health benefits, side effects, and risks have been considered in formulating the recommendations.

Rating: 7

The health benefits, side effects, and risks have been considered in formulating the recommendations.

##### 12. There is an explicit link between the recommendations and the supporting evidence.

Rating: 7

There is an explicit link between the recommendations and the supporting evidence.

##### 13. The guideline has been externally reviewed by experts prior to its publication.

Rating: 7

A report of the consultation and peer review comments and responses is available in the supporting material section for this guideline on the SIGN website.

##### 14. A procedure for updating the guideline is provided.

Rating: 6

A procedure for updating the guideline is provided but not so clear in the interim period or criteria for updating.

---

#### 4. Clarity of Presentation

##### 15. The recommendations are specific and unambiguous.

Rating: 7

Recommendations provide concrete and precise descriptions of which option is appropriate in which situation and in what population group, as informed by the body of evidence.

##### 16. The different options for management of the condition or health issue are clearly presented.

Rating: 7

The different options for management of the condition or health issue are clearly presented.

##### 17. Key recommendations are easily identifiable.

Rating: 7

Key recommendations are easily identifiable in the Whole of chapter 2.

---

### 5. Applicability

#### 18. The guideline describes facilitators and barriers to its application.

Rating: 7

The guideline describes the facilitators and barriers to its application.

#### 19. The guideline provides advice and/or tools on how the recommendations can be put into practice.

Rating: 7

The guideline provides tools on how the recommendations can be put into practice.

#### 20. The potential resource implications of applying the recommendations have been considered.

Rating: 7

The potential resource implications of applying the recommendations and economic costs have been considered.

#### 21. The guideline presents monitoring and/or auditing criteria.

Rating: 7

The guideline presents clear monitoring criteria for each recommendation.

---

### 6. Editorial Independence

#### 22. The views of the funding body have not influenced the content of the guideline.

Rating: 6

The views of the funding body have not influenced the content of the guideline, but not in a clear way.

#### 23. Competing interests of guideline development group members have been recorded and addressed.

Rating: 6

Competing interests of guideline development group members have been recorded and addressed, but not in a clear way.

---

Created online at www.agreetrust.org 26 October 2022

### A critical appraisal of:
### Primary & Secondary Prevention of Cardiovascular Disease / 2017
### using the AGREE II Instrument

Created with the AGREE II Online Guideline Appraisal Tool.

No endorsement of the content of this document by the AGREE Research Trust should be implied.

Appraiser: Mohammed Abd ElFattah Darwesh Badawy

Date: 30 October 2022

URL of this appraisal: http://www.agreetrust.org/appraisal/97115

Guideline URL:

---

#### Overall Assessment

Title: Primary & Secondary Prevention of Cardiovascular Disease / 2017

Overall quality of this guideline: 7/7

Guideline recommended for use? Yes.

Notes:

Perfect guideline for CVD Primary & Secondary Prevention for Malaysian population.

| Domain | Total |
|---|---|
| 1. Scope and Purpose | 21 |
| 2. Stakeholder Involvement | 21 |
| 3. Rigour of Development | 55 |
| 4. Clarity of Presentation | 21 |
| 5. Applicability | 28 |
| 6. Editorial Independence | 14 |

---

### 1. Scope and Purpose

#### 1. The overall objective(s) of the guideline is (are) specifically described.

Rating: 7

The overall objectives of the guideline are specifically described in \"STATEMENT OF INTENT\".

#### 2. The health question(s) covered by the guideline is (are) specifically described.

Rating: 7

The health question covered by the guideline is specifically described.

#### 3. The population (patients, public, etc.) to whom the guideline is meant to apply is specifically described.

Rating: 7

The population to whom the guideline is meant to apply is specifically described.

---

### 2. Stakeholder Involvement

#### 4. The guideline development group includes individuals from all relevant professional groups.

Rating: 7

\"MEMBERS OF THE EXPERT PANEL\" includes individuals from all relevant professional groups.

##### 5. The views and preferences of the target population (patients, public, etc.) have been sought.

Rating: 7

The views and preferences of the target population (patients, public, etc.) have been sought.

##### 6. The target users of the guideline are clearly defined.

Rating: 7

The target users of the guideline are clearly defined.

---

#### 3. Rigour of Development

##### 7. Systematic methods were used to search for evidence.

Rating: 7

Systematic methods were used to search for evidence.

##### 8. The criteria for selecting the evidence are clearly described.

Rating: 7

The criteria for selecting the evidence are clearly described.

##### 9. The strengths and limitations of the body of evidence are clearly described.

Rating: 7

The strengths and limitations of the body of evidence are clearly described.

##### 10. The methods for formulating the recommendations are clearly described.

Rating: 6

The methods for formulating the recommendations are clearly described.

##### 11. The health benefits, side effects, and risks have been considered in formulating the recommendations.

Rating: 7

The health benefits, side effects, and risks have been considered in formulating the recommendations.

##### 12. There is an explicit link between the recommendations and the supporting evidence.

Rating: 7

There is an explicit link between the recommendations and the supporting evidence.

##### 13. The guideline has been externally reviewed by experts prior to its publication.

Rating: 7

The guideline has been externally reviewed by experts prior to its publication and \" EXTERNAL REVIEWERS\" are listed.

##### 14. A procedure for updating the guideline is provided.

Rating: 7

A procedure for updating the guideline is provided to be updated every 5 years.

---

#### 4. Clarity of Presentation

##### 15. The recommendations are specific and unambiguous.

Rating: 7

The recommendations are specific and unambiguous.

##### 16. The different options for management of the condition or health issue are clearly presented.

Rating: 7

The different options for management of the condition or health issue are clearly presented.

##### 17. Key recommendations are easily identifiable.

Rating: 7

Key recommendations are easily identifiable.

---

### 5. Applicability

##### 18. The guideline describes facilitators and barriers to its application.

Rating: 7

The guideline describes facilitators and barriers to its application.

##### 19. The guideline provides advice and/or tools on how the recommendations can be put into practice.

Rating: 7

The guideline provides advice and tools on how the recommendations can be implemented.

##### 20. The potential resource implications of applying the recommendations have been considered.

Rating: 7

The potential resource implications of applying the recommendations have been considered.

##### 21. The guideline presents monitoring and/or auditing criteria.

Rating: 7

The guideline presents monitoring criteria.

---

### 6. Editorial Independence

##### 22. The views of the funding body have not influenced the content of the guideline.

Rating: 7

The views of the funding body have not influenced the content of the guideline.

##### 23. Competing interests of guideline development group members have been recorded and addressed.

Rating: 7

Competing interests of guideline development group members have been recorded and addressed.

---

Created online at www.agreetrust.org 30 October 2022

### A critical appraisal of:
### Canadian Cardiovascular Harmonized National Guidelines Endeavour (C-CHANGE) guideline for the prevention and management of cardiovascular disease in primary care / 2018
### using the AGREE II Instrument

Created with the AGREE II Online Guideline Appraisal Tool.

No endorsement of the content of this document by the AGREE Research Trust should be implied.

Appraiser: Mohammed Abd ElFattah Darwesh Badawy

Date: 26 October 2022

URL of this appraisal: http://www.agreetrust.org/appraisal/97118

Guideline URL:

### Overall Assessment

Title: Canadian Cardiovascular Harmonized National Guidelines Endeavour (C-CHANGE) guideline for the prevention and management of cardiovascular disease in primary care / 2018

Overall quality of this guideline: 7/7

Guideline recommended for use? Yes.

| Domain | Total |
|---|---|
| 1. Scope and Purpose | 21 |
| 2. Stakeholder Involvement | 19 |
| 3. Rigour of Development | 56 |
| 4. Clarity of Presentation | 20 |
| 5. Applicability | 25 |
| 6. Editorial Independence | 14 |

### 1. Scope and Purpose

#### 1. The overall objective(s) of the guideline is (are) specifically described.

Rating: 7

The overall objective of the guideline is specifically described.

#### 2. The health question(s) covered by the guideline is (are) specifically described.

Rating: 7

The health question covered by the guideline is specifically described.

#### 3. The population (patients, public, etc.) to whom the guideline is meant to apply is specifically described.

Rating: 7

The population (patients, public, etc.) to whom the guideline is meant to apply is specifically described.

### 2. Stakeholder Involvement

#### 4. The guideline development group includes individuals from all relevant professional groups.

Rating: 7

The guideline development group includes individuals from all relevant professional groups.

#### 5. The views and preferences of the target population (patients, public, etc.) have been sought.

Rating: 6

The views and preferences of the target population (patients, public, etc.) have been sought, but not many details are described.

#### 6. The target users of the guideline are clearly defined.

Rating: 6

The target users of the guideline are clearly defined.

---

### 3. Rigour of Development

#### 7. Systematic methods were used to search for evidence.

Rating: 7

Systematic methods were used to search for evidence.

#### 8. The criteria for selecting the evidence are clearly described.

Rating: 7

The criteria for selecting the evidence are clearly described.

#### 9. The strengths and limitations of the body of evidence are clearly described.

Rating: 7

The strengths and limitations of the body of evidence are clearly described.

#### 10. The methods for formulating the recommendations are clearly described.

Rating: 7

The methods for formulating the recommendations are clearly described.

##### 11. The health benefits, side effects, and risks have been considered in formulating the recommendations.

Rating: 7

The health benefits, side effects, and risks have been considered in formulating the recommendations.

##### 12. There is an explicit link between the recommendations and the supporting evidence.

Rating: 7

There is an explicit link between the recommendations and the supporting evidence.

##### 13. The guideline has been externally reviewed by experts prior to its publication.

Rating: 7

The guideline has been externally reviewed by experts prior to its publication.

##### 14. A procedure for updating the guideline is provided.

Rating: 7

A procedure for updating the guideline is provided.

---

#### 4. Clarity of Presentation

##### 15. The recommendations are specific and unambiguous.

Rating: 7

The recommendations are specific and unambiguous.

##### 16. The different options for management of the condition or health issue are clearly presented.

Rating: 6

The different options for the management of the condition or health issue are clearly presented.

##### 17. Key recommendations are easily identifiable.

Rating: 7

Key recommendations are easily identifiable.

---

### 5. Applicability

#### 18. The guideline describes facilitators and barriers to its application.

Rating: 6

The guideline describes the facilitators and barriers to its application.

#### 19. The guideline provides advice and/or tools on how the recommendations can be put into practice.

Rating: 7

The guideline provides advice and tools on how the recommendations can be put into practice.

#### 20. The potential resource implications of applying the recommendations have been considered.

Rating: 6

The potential resource implications of applying the recommendations have been considered.

#### 21. The guideline presents monitoring and/or auditing criteria.

Rating: 6

The guideline presents monitoring and auditing criteria.

---

### 6. Editorial Independence

#### 22. The views of the funding body have not influenced the content of the guideline.

Rating: 7

The views of the funding body have not influenced the content of the guideline.

#### 23. Competing interests of guideline development group members have been recorded and addressed.

Rating: 7

Competing interests of guideline development group members have been recorded and addressed.

---

Created online at www.agreetrust.org 26 October 2022

### A critical appraisal of: Cardiovascular Disease Risk Assessment and Management for Primary Care / 2018 using the AGREE II Instrument

Created with the AGREE II Online Guideline Appraisal Tool.

No endorsement of the content of this document by the AGREE Research Trust should be implied.

Appraiser: Mohammed Abd ElFattah Darwesh Badawy

Date: 30 October 2022

URL of this appraisal: http://www.agreetrust.org/appraisal/97280

Guideline URL:

---

#### Overall Assessment

Title: Cardiovascular Disease Risk Assessment and Management for Primary Care / 2018

Overall quality of this guideline: 5/7

Guideline recommended for use? Yes with modifications.

| Domain | Total |
|---|---|
| 1. Scope and Purpose | 18 |
| 2. Stakeholder Involvement | 17 |
| 3. Rigour of Development | 46 |
| 4. Clarity of Presentation | 18 |
| 5. Applicability | 24 |
| 6. Editorial Independence | 10 |

### 1. Scope and Purpose

#### 1. The overall objective(s) of the guideline is (are) specifically described.

Rating: 6

The overall objective of the guideline is specifically described

#### 2. The health question(s) covered by the guideline is (are) specifically described.

Rating: 6

The health question covered by the guideline is specifically described.

#### 3. The population (patients, public, etc.) to whom the guideline is meant to apply is specifically described.

Rating: 6

The population to whom the guideline is meant to apply is specifically described.

### 2. Stakeholder Involvement

#### 4. The guideline development group includes individuals from all relevant professional groups.

Rating: 5

The professionals who were involved in the development process are included but without any details of their roles.

##### 5. The views and preferences of the target population (patients, public, etc.) have been sought.

Rating: 5

The views and preferences of the target population are not well-informed in the guideline.

##### 6. The target users of the guideline are clearly defined.

Rating: 7

The target users of the guideline are clearly defined.

---

#### 3. Rigour of Development

##### 7. Systematic methods were used to search for evidence.

Rating: 7

Systematic methods were used to search for evidence.

##### 8. The criteria for selecting the evidence are clearly described.

Rating: 7

The criteria for selecting the evidence are clearly described.

##### 9. The strengths and limitations of the body of evidence are clearly described.

Rating: 5

The strengths and limitations of the body of evidence are not clearly described.

##### 10. The methods for formulating the recommendations are clearly described.

Rating: 5

The methods for formulating the recommendations are not clearly described.

##### 11. The health benefits, side effects, and risks have been considered in formulating the recommendations.

Rating: 7

The health benefits, side effects, and risks have been considered in formulating the recommendations.

##### 12. There is an explicit link between the recommendations and the supporting evidence.

Rating: 4

The link between the recommendations and the supporting evidence is not evident.

##### 13. The guideline has been externally reviewed by experts prior to its publication.

Rating: 5

The guideline has been externally reviewed by experts before its publication.

##### 14. A procedure for updating the guideline is provided.

Rating: 6

A procedure for updating the guideline is not clearly provided.

---

#### 4. Clarity of Presentation

##### 15. The recommendations are specific and unambiguous.

Rating: 6

The recommendations are specific and unambiguous.

##### 16. The different options for management of the condition or health issue are clearly presented.

Rating: 6

The different options for the management of the condition or health issue are clearly presented.

##### 17. Key recommendations are easily identifiable.

Rating: 6

Key recommendations are easily identifiable.

---

#### 5. Applicability

##### 18. The guideline describes facilitators and barriers to its application.

Rating: 6

The guideline describes the facilitators and barriers to its application.

##### 19. The guideline provides advice and/or tools on how the recommendations can be put into practice.

Rating: 5

The guideline provides advice on how the recommendations can be put into practice.

##### 20. The potential resource implications of applying the recommendations have been considered.

Rating: 6

The potential resource implications of applying the recommendations have been considered.

##### 21. The guideline presents monitoring and/or auditing criteria.

Rating: 7

The guideline presents monitoring and auditing criteria.

---

#### 6. Editorial Independence

##### 22. The views of the funding body have not influenced the content of the guideline.

Rating: 5

A statement that the funding body did not influence the content of the guideline is not evident.

##### 23. Competing interests of guideline development group members have been recorded and addressed.

Rating: 5

The competing interests of guideline development group members have not been well-recorded.

---

Created online at www.agreetrust.org 30 October 2022

### A critical appraisal of:
### AHA/ACC Guideline on the Primary Prevention of Cardiovascular Disease / 2019
### using the AGREE II Instrument

Created with the AGREE II Online Guideline Appraisal Tool.

No endorsement of the content of this document by the AGREE Research Trust should be implied.


Appraiser: Mohammed Abd ElFattah Darwesh Badawy

Date: 31 October 2022


URL of this appraisal: http://www.agreetrust.org/appraisal/97279

Guideline URL:

### Overall Assessment

Title: AHA/ACC Guideline on the Primary Prevention of Cardiovascular Disease / 2019

Overall quality of this guideline: 6/7

Guideline recommended for use? Yes with modifications.

| Domain | Total |
|---|---|
| 1. Scope and Purpose | 19 |
| 2. Stakeholder Involvement | 19 |
| 3. Rigour of Development | 46 |
| 4. Clarity of Presentation | 20 |
| 5. Applicability | 26 |
| 6. Editorial Independence | 8 |

### 1. Scope and Purpose

#### 1. The overall objective(s) of the guideline is (are) specifically described.

Rating: 7

The overall objectives of the guideline are specifically described under the section \"Scope of the Guideline.\"

#### 2. The health question(s) covered by the guideline is (are) specifically described.

Rating: 6

The health questions covered by the guideline are specifically described.

#### 3. The population (patients, public, etc.) to whom the guideline is meant to apply is specifically described.

Rating: 6

The population to whom the guideline is meant to apply is specifically described.

### 2. Stakeholder Involvement

##### 4. The guideline development group includes individuals from all relevant professional groups.

Rating: 7

The guideline development group includes individuals from all relevant professional groups with a detailed description of the member’s role in the guideline development group.

##### 5. The views and preferences of the target population (patients, public, etc.) have been sought.

Rating: 6

The views and preferences of the target population have been sought.

##### 6. The target users of the guideline are clearly defined.

Rating: 6

The target users of the guideline are clearly defined.

---

#### 3. Rigour of Development

##### 7. Systematic methods were used to search for evidence.

Rating: 7

Systematic methods were used to search for evidence.

##### 8. The criteria for selecting the evidence are clearly described.

Rating: 6

The criteria for selecting the evidence are clearly described.

##### 9. The strengths and limitations of the body of evidence are clearly described.

Rating: 7

The strengths and limitations of the body of evidence are clearly described.

##### 10. The methods for formulating the recommendations are clearly described.

Rating: 3

The methods for formulating the recommendations are not clearly described.

##### 11. The health benefits, side effects, and risks have been considered in formulating the recommendations.

Rating: 6

The health benefits, side effects, and risks have been considered in formulating the recommendations.

##### 12. There is an explicit link between the recommendations and the supporting evidence.

Rating: 7

There is an explicit link between the recommendations and the supporting evidence.

##### 13. The guideline has been externally reviewed by experts prior to its publication.

Rating: 7

The guideline has been externally reviewed by experts prior to its publication.

##### 14. A procedure for updating the guideline is provided.

Rating: 3

A procedure for updating the guideline is not clearly provided.

---

#### 4. Clarity of Presentation

##### 15. The recommendations are specific and unambiguous.

Rating: 6

The recommendations are specific and unambiguous.

##### 16. The different options for management of the condition or health issue are clearly presented.

Rating: 7

The different options for the management of the condition or health issue are clearly presented.

##### 17. Key recommendations are easily identifiable.

Rating: 7

Key recommendations are easily identifiable.

---

### 5. Applicability

#### 18. The guideline describes facilitators and barriers to its application.

Rating: 7

The guideline describes facilitators and barriers to its application.

#### 19. The guideline provides advice and/or tools on how the recommendations can be put into practice.

Rating: 6

The guideline provides advice on how the recommendations can be put into practice.

#### 20. The potential resource implications of applying the recommendations have been considered.

Rating: 7

The potential resource implications of applying the recommendations have been considered.

#### 21. The guideline presents monitoring and/or auditing criteria.

Rating: 6

The guideline presents monitoring and auditing criteria.

---

### 6. Editorial Independence

#### 22. The views of the funding body have not influenced the content of the guideline.

Rating: 3

There is no clear statement that the funding body did not influence the content of the guideline

#### 23. Competing interests of guideline development group members have been recorded and addressed.

Rating: 5

The competing interests of guideline development group members are not well-recorded.

Created online at www.agreetrust.org 31 October 2022

### A critical appraisal of:
### National Health Screening Guideline on Noncommunicable Diseases (NCDs) / 2019
### using the AGREE II Instrument

Created with the AGREE II Online Guideline Appraisal Tool.

No endorsement of the content of this document by the AGREE Research Trust should be implied.

Appraiser: Mohammed Abd ElFattah Darwesh Badawy

Date: 29 October 2022

URL of this appraisal: http://www.agreetrust.org/appraisal/97278

Guideline URL:

### Overall Assessment

Title: National Health Screening Guideline on Noncommunicable Diseases (NCDs) / 2019

Overall quality of this guideline: 5/7

Guideline recommended for use? Yes with modifications.

| Domain | Total |
|---|---|
| 1. Scope and Purpose | 16 |
| 2. Stakeholder Involvement | 16 |
| 3. Rigour of Development | 28 |
| 4. Clarity of Presentation | 19 |
| 5. Applicability | 15 |
| 6. Editorial Independence | 8 |

### 1. Scope and Purpose

##### 1. The overall objective(s) of the guideline is (are) specifically described.

Rating: 5

The overall objective of the guideline is described at the beginning of the guideline.

##### 2. The health question(s) covered by the guideline is (are) specifically described.

Rating: 5

The health questions covered by the guideline are described.

##### 3. The population (patients, public, etc.) to whom the guideline is meant to apply is specifically described.

Rating: 6

The population (patients, public, etc.) to whom the guideline is meant to apply is specifically described

### 2. Stakeholder Involvement

##### 4. The guideline development group includes individuals from all relevant professional groups.

Rating: 6

The guideline development group includes individuals from all relevant professional groups and is described briefly in the guideline.

##### 5. The views and preferences of the target population (patients, public, etc.) have been sought.

Rating: 3

Information about the target population\'s experiences and expectations of health care are not clearly informed.

##### 6. The target users of the guideline are clearly defined.

Rating: 7

The target users of the guideline are clearly defined.

---

#### 3. Rigour of Development

##### 7. Systematic methods were used to search for evidence.

Rating: 5

Systematic methods were used to search for evidence, but the details are not included.

##### 8. The criteria for selecting the evidence are clearly described.

Rating: 3

The criteria for selecting the evidence are not clearly described.

##### 9. The strengths and limitations of the body of evidence are clearly described.

Rating: 3

The strengths and limitations of the body of evidence are not clearly described.

##### 10. The methods for formulating the recommendations are clearly described.

Rating: 3

The methods for formulating the recommendations are not clearly described

##### 11. The health benefits, side effects, and risks have been considered in formulating the recommendations.

Rating: 5

The health benefits, side effects, and risks have been considered in formulating the recommendations.

##### 12. There is an explicit link between the recommendations and the supporting evidence.

Rating: 3

There is no clear, explicit link between the recommendations and the supporting evidence.

##### 13. The guideline has been externally reviewed by experts prior to its publication.

Rating: 3

The external review is not evident in this guideline.

##### 14. A procedure for updating the guideline is provided.

Rating: 3

A procedure for updating the guideline is not clearly provided.

---

#### 4. Clarity of Presentation

##### 15. The recommendations are specific and unambiguous.

Rating: 7

The recommendations are specific and unambiguous.

##### 16. The different options for management of the condition or health issue are clearly presented.

Rating: 5

The different options for the management of the condition or health issue are presented but not in a clear way.

##### 17. Key recommendations are easily identifiable.

Rating: 7

Key recommendations are easily identifiable

### 5. Applicability

##### 18. The guideline describes facilitators and barriers to its application.

Rating: 3

The guideline does not describe facilitators and barriers to its application.

##### 19. The guideline provides advice and/or tools on how the recommendations can be put into practice.

Rating: 3

The guideline does not provide advice or tools on how the recommendations can be put into practice.

##### 20. The potential resource implications of applying the recommendations have been considered.

Rating: 6

The potential resource implications of applying the recommendations have been considered.

##### 21. The guideline presents monitoring and/or auditing criteria.

Rating: 3

The guideline does not present monitoring or auditing criteria.

### 6. Editorial Independence

##### 22. The views of the funding body have not influenced the content of the guideline.

Rating: 3

The views of the funding body are not clear in the guideline.

##### 23. Competing interests of guideline development group members have been recorded and addressed.

Rating: 5

The competing interests of guideline development group members have been fairly addressed.

Created online at www.agreetrust.org 29 October 2022

### A critical appraisal of: ESC Guidelines on cardiovascular disease prevention in clinical practice / 2021 using the AGREE II Instrument

Created with the AGREE II Online Guideline Appraisal Tool.

No endorsement of the content of this document by the AGREE Research Trust should be implied.

Appraiser: Mohammed Abd ElFattah Darwesh Badawy

Date: 28 October 2022

URL of this appraisal: http://www.agreetrust.org/appraisal/97268

Guideline URL:

---

#### Overall Assessment

Title: ESC Guidelines on cardiovascular disease prevention in clinical practice / 2021

Overall quality of this guideline: 7/7

Guideline recommended for use? Yes.

| Domain | Total |
| --- | --- |
| 1. Scope and Purpose | 18 |
| 2. Stakeholder Involvement | 21 |
| 3. Rigour of Development | 48 |
| 4. Clarity of Presentation | 20 |
| 5. Applicability | 26 |
| 6. Editorial Independence | 14 |

---

### 1. Scope and Purpose

##### 1. The overall objective(s) of the guideline is (are) specifically described.

Rating: 6

The overall objective of the guideline is specifically described but not easy to find in the text.

##### 2. The health question(s) covered by the guideline is (are) specifically described.

Rating: 6

The health question covered by the guideline is specifically described but not easy to find in the text.

##### 3. The population (patients, public, etc.) to whom the guideline is meant to apply is specifically described.

Rating: 6

The population to whom the guideline is meant to apply is specifically described.

---

### 2. Stakeholder Involvement

##### 4. The guideline development group includes individuals from all relevant professional groups.

Rating: 7

The guideline development group includes individuals from all relevant professional groups.

##### 5. The views and preferences of the target population (patients, public, etc.) have been sought.

Rating: 7

The views and preferences of the target population have been sought.

##### 6. The target users of the guideline are clearly defined.

Rating: 7

The target users of the guideline are clearly defined.

---

#### 3. Rigour of Development

##### 7. Systematic methods were used to search for evidence.

Rating: 5

Details of the strategy used to search for evidence are not clearly provided.

##### 8. The criteria for selecting the evidence are clearly described.

Rating: 5

The criteria for selecting the evidence are not clearly described.

##### 9. The strengths and limitations of the body of evidence are clearly described.

Rating: 7

The strengths and limitations of the body of evidence are clearly described.

##### 10. The methods for formulating the recommendations are clearly described.

Rating: 5

The methods for formulating the recommendations are not clearly described.

##### 11. The health benefits, side effects, and risks have been considered in formulating the recommendations.

Rating: 7

The health benefits, side effects, and risks have been considered in formulating the recommendations.

##### 12. There is an explicit link between the recommendations and the supporting evidence.

Rating: 7

There is an explicit link between the recommendations and the supporting evidence.

##### 13. The guideline has been externally reviewed by experts prior to its publication.

Rating: 7

The guideline has been externally reviewed by experts prior to its publication.

##### 14. A procedure for updating the guideline is provided.

Rating: 5

A procedure for updating the guideline is not clearly provided.

---

#### 4. Clarity of Presentation

##### 15. The recommendations are specific and unambiguous.

Rating: 7

The recommendations are specific and unambiguous.

##### 16. The different options for management of the condition or health issue are clearly presented.

Rating: 6

The different options for management of the condition or health issue are clearly presented.

##### 17. Key recommendations are easily identifiable.

Rating: 7

Key recommendations are easily identifiable.

---

#### 5. Applicability

##### 18. The guideline describes facilitators and barriers to its application.

Rating: 7

The guideline describes the facilitators and barriers to its application.

##### 19. The guideline provides advice and/or tools on how the recommendations can be put into practice.

Rating: 6

The guideline provides advice on how the recommendations can be put into practice.

##### 20. The potential resource implications of applying the recommendations have been considered.

Rating: 6

The potential resource implications of applying the recommendations have been considered.

##### 21. The guideline presents monitoring and/or auditing criteria.

Rating: 7

The guideline presents monitoring and auditing criteria.

---

#### 6. Editorial Independence

##### 22. The views of the funding body have not influenced the content of the guideline.

Rating: 7

The views of the funding body have not influenced the content of the guideline.

##### 23. Competing interests of guideline development group members have been recorded and addressed.

Rating: 7

Competing interests of guideline development group members have been recorded and addressed.

---

Created online at www.agreetrust.org 28 October 2022
